# Type 1 diabetes risk and severity after SARS-CoV-2 infection or vaccination

**DOI:** 10.1101/2024.07.03.24309894

**Authors:** Lars C. Stene, Paz Lopez-Doriga Ruiz, Rickard Ljung, Håkon Bøås, Hanne L. Gulseth, Nicklas Pihlström, Anders Sundström, Björn Zethelius, Ketil Størdal, Osman Gani, Nicolai A. Lund-Blix, Torild Skrivarhaug, German Tapia

## Abstract

**Aim:** To clarify whether SARS-CoV-2 infection or vaccination contribute to risk of type 1 diabetes or more severe diabetes onset in children and young adults.

**Methods:** We analysed cohorts of population-wide registries of young individuals from Norway (N=1,986,970) and Sweden (N=2,100,188). We used regression models to estimate adjusted rate ratios (aRR), treating exposures as time-varying, starting 30 days after registered SARS-CoV-2 positive test or vaccination.

**Findings:** Pooled results from Norway and Sweden and age-groups 12-17 and 18-29 years showed no significant increase in type 1 diabetes after documented infections (aRR 1.06, 95%CI:0.77-1.45). There was moderate heterogeneity, with a suggestive increased risk among children in Norway after infection. Pooled results for Norway and Sweden and age-groups 12-17 years and 18-29 years showed no significant association between SARS-CoV-2 vaccination and risk of type 1 diabetes (aRR 1.09, 95%CI: 0.81, 1.48). There was significant heterogeneity, primarily driven by a positive association among children and an inverse association in young adults in Sweden. While the type 1 diabetes incidence increased and diabetes ketoacidosis decreased over time during 2016-2023, no significant break in time-trends were seen after March 2020 for HbA1c, risk or severity of diabetic ketoacidosis, or islet autoantibodies, at diagnosis of type 1 diabetes.

**Interpretation:** Taken together, these results do not indicate any consistent, large effects of SARS-CoV-2 infection or -vaccination on risk of type 1 diabetes or severity at disease onset. Suggestive associations in sub-groups should be investigated further in other studies.

**Funding:** The work was done as part of regular work at the institutions where the authors had their primary affiliation, and no specific funding was obtained for these studies.

## Introduction

Severe Acute Respiratory Syndrome Coronavirus-2 (SARS-CoV-2) infection has been associated with a wide variety of post-acute conditions, affecting most organ systems.^1^ Symptoms range from none in the majority to fatal in a minority.^2^

Type 1 diabetes is an immune-mediated disease with increasing incidence globally, and the Nordic countries having high incidence rates compared to most other regions.^3^ Few environmental factors have been unequivocally implicated in the aetiology, but common viral infections have been suspected.^4–6^ Enteroviruses have been most frequently studied, and respiratory infections have also been associated with subsequent risk of type 1 diabetes.^7–9^ It is therefore not surprising that a potential link between SARS-CoV-2 and incident type 1 diabetes has been hypothesized.^10^ The evidence for this link consists of increases in type 1 diabetes incidence after the start of the corona pandemic, case-reports of SARS-CoV-2 in new onset patients with type 1 diabetes and various in vitro studies investigating the possible presence of SARS-CoV-2 virus in human pancreatic islets and beta-cells and their expression of viral entry receptors.^11–13^ The latter has yielded mixed results depending on experimental conditions.^14^ Potential indirect effects of the pandemic including reduced occurrence of other viral infections may explain some of the changing incidence or seasonality of type 1 diabetes after the pandemic started. It has been hypothesized that the lockdown led to delayed diagnosis with consequent increase in the proportion of new onset cases who had diabetic ketoacidosis.^11,15^

Large prospective studies investigating the risk of type 1 diabetes in young individuals after testing positive for SARS-CoV-2 compared to those without a positive test have yielded mixed results. Whole population registry-based studies from Scotland and Denmark did not find significantly increased risk of type 1 diabetes in the post-acute follow-up period after SARS-CoV-2 positive PCR in children or young adults,^16–18^ whereas other studies using claims databases have reported approximately 1.5-fold increased risk of type 1 diabetes.^19–21^ Most previous studies did not adjust analyses for SARS-CoV-2 vaccination. Finally, it has been speculated that vaccines can induce autoimmune diseases. A few case reports of type 1 diabetes after SARS-CoV-2 vaccines have been reported,^22^ but we are aware of only one published cohort study.^16^

Norway and Sweden are neighbouring countries with universally available health care system free of charge to citizens, nationwide registries, high vaccine coverage, and high SARS-CoV-2 testing rates August 2020-February 2022. During the pandemic Sweden took a less restrictive approach to mitigate the pandemic in the early phase, compared to Norway. Using individual level nationwide registry linkages in Norway and Sweden, we aimed to investigate the incidence trends of type 1 diabetes before and during the pandemic, clinical characteristics at onset suggestive of delayed diagnosis, and type 1 diabetes incidence after SARS-CoV-2 infection or after vaccination.

## Methods

### Participants and study design

We used three population-based cohorts, the Norwegian Childhood Diabetes Registry (NCDR), and two sets of individual-level linked total population registers from Norway and Sweden (Figure 1). Detailed time trends including seasonality of incidence and severity at onset (blood pH, bicarbonate, glycated haemoglobin and presence of islet autoantibodies) in all children under 18 years of age with newly diagnosed type 1 diabetes in Norway from 2016 to 2023 were studied using NCDR-data. The total populations aged 0-29 years (Norway) and 12-29 years (Sweden) in 2020 were used to study the association between SARS-CoV-2 infection (based on PCR or antigen tests) or vaccination and subsequent risk of type 1 diabetes up to 2022.^23–25^ The registers contain data on diagnosis codes from inpatient and outpatient care, prescribed blood glucose lowering medications, SARS-CoV-2 positive and negative test results, SARS-CoV-2 vaccination dates and products used, and socio-demographic variables (disease and medication codes listed in Supplemental Table S1). The studies were approved by the ethics committees in the respective countries. See Supplemental Appendix Methods for further details on data sources and ethics approvals.

**Figure 1.**
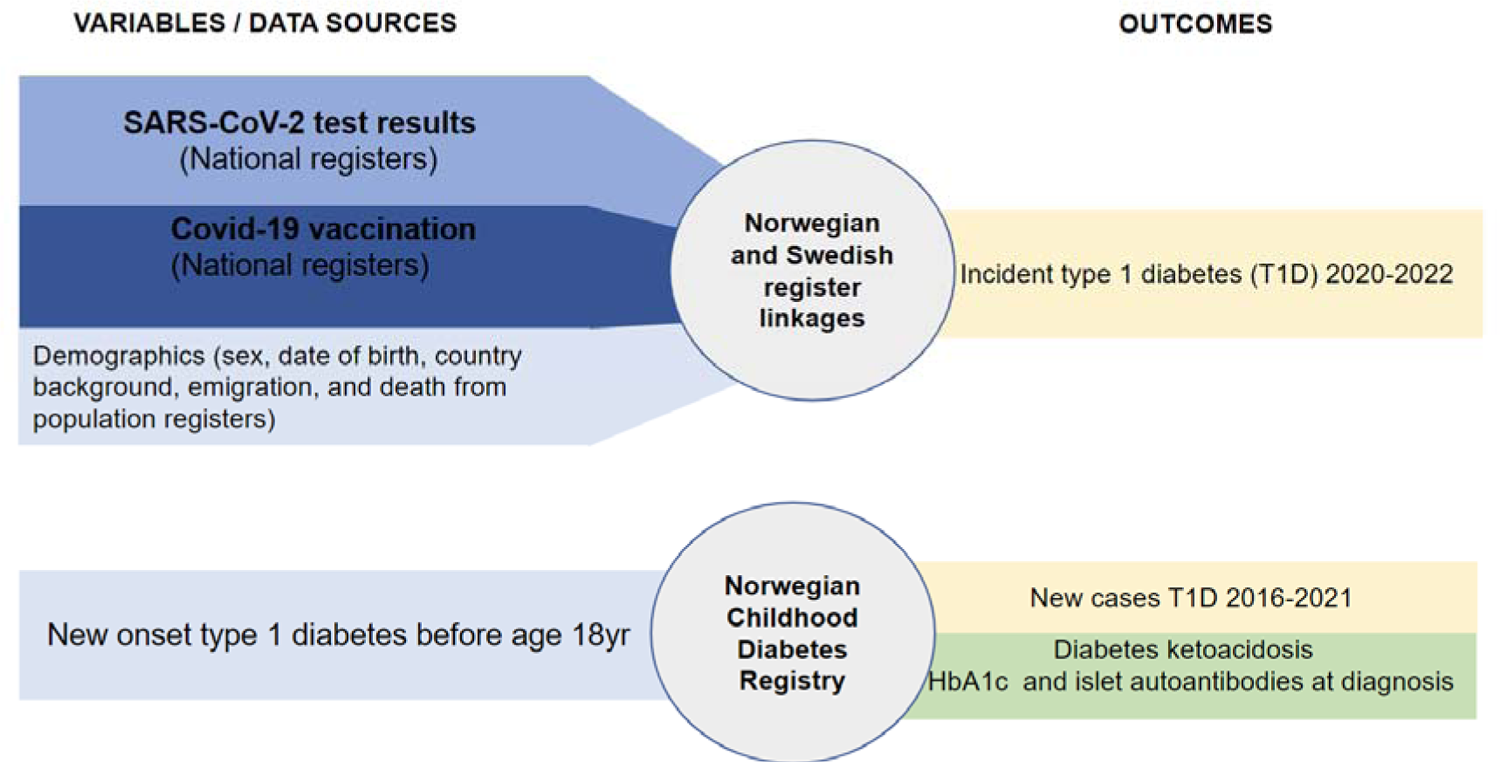
Overview of study design and data sources. Data sources, exposures and outcomes in three independent population-based cohort studies.

### Outcome: Type 1 diabetes and clinical characteristics at diagnosis

For the total population cohorts in Norway and Sweden, the endpoint was diagnosis of incident type 1 diabetes. This was defined as a type 1 diabetes diagnosis code in the patient registry plus insulin use. To avoid including prevalent cases, we excluded individuals with a previous diabetes diagnosis or previous use of any glucose-lowering medications during 2017-2019 in Norway and 2015-2019 in Sweden (see Supplemental Methods for detailed algorithm).

For the NCDR, the date of diagnosis is defined as the day of the first insulin injection. Diabetic ketoacidosis (DKA) was defined as blood pH <7.3 or serum bicarbonate <15 mmol/L. Glycated haemoglobin (HbA1c) was measured at admission at the paediatric departments and recorded in the NCDR. Similarly, we included data on positivity for islet autoantibodies at diagnosis of type 1 diabetes (see Supplemental Methods for further details and references).

### Exposures: SARS-CoV-2 infection and SARS-CoV-2 vaccination

SARS-CoV-2 infection was defined as having a registered positive PCR or rapid antigen test. Both SARS-CoV-2 infection and vaccination were modelled as time-varying exposures counted from the 31^st^ day after the first registered positive test or vaccination. We also did additional robustness analyses by explicitly modelling also the second vaccination dose. Essentially all vaccines given in these age-groups were mRNA vaccines (additional details in Supplemental Methods).

### Statistical analysis

Incidence rates (number of incident cases diagnosed with type 1 diabetes per 100,000 person-years) for SARS-CoV-2 infection- or vaccination exposed and non-exposed groups were estimated using the register linkages from Norway and Sweden, separately for children and young adults. We estimated adjusted rate ratios (aRR) with 95% confidence intervals to assess the potential association between SARS-CoV-2 infection or vaccination and type 1 diabetes using Poisson regression on the Swedish data and Cox regression on the Norwegian data (calendar time was the primary time-variable). Analyses were adjusted for age, sex and Nordic or non-Nordic country background. Additional adjustments for coeliac disease, family income, crowding, household size, urban/rural residence and geographical regions were done for Norwegian analyses. In addition to full cohort analyses in both countries, we also analysed Norwegian data according to the test-negative design including only PCR-tested individuals.^26^ Robustness analyses were done to comparing cohort and test-negative analyses and results in age-groups. See Supplemental Methods for details on statistical analysis.

Incidence and seasonality of new onset type 1 diabetes before and during the pandemic was modelled as the monthly number of newly diagnosed cases, using negative binomial regression models with restricted cubic splines for calendar time. We used interrupted time-series modelling to assess potential impacts of the first lockdown 12 March 2020. Similar analyses were done using logistic models for binary outcomes, and linear regression for continuous outcome variables. Changes in seasonality of type 1 diabetes incidence after the first lockdown was tested using the cosinor model (see Supplemental Methods for further detail).

### Role of the funding source

The work was done as part of regular work at the institutions where the authors had their primary affiliation, and no specific funding was obtained for these studies.

## Results

An overview of study designs and data sources are shown in Figure 1. A total of 1,986,970 Norwegian residents aged 0-29 years during 2020-2022 and 2,100,188 Swedish residents aged 12-29 years were analysed, of which 1,307 developed type 1 diabetes during follow-up in Norway and 1,021 in Sweden. Approximately 70% had both a mother and father of Nordic origin in both Norway and Sweden (characteristics in Supplemental Table S2). In addition, time-trends in incidence and severity at onset of type 1 diabetes were studied in 2913 Norwegians aged under 18 years 2016-2023.

### SARS-CoV-2 infection and subsequent risk of type 1 diabetes

SARS-CoV-2 infection frequency was low during spring 2020 before it increased in smaller waves with the large majority occurring after November 2021 coinciding with the introduction of the omicron variant (Supplemental Figure S1). The time from infection to diagnosis of type 1 diabetes showed a relatively wide distribution with no apparent clustering apart from the highest number of cases in the first few days after infection (Supplemental Figure S2 and S3a).

Pooled results from Norway and Sweden and age-groups 12-17 and 18-29 years showed no significant increase in type 1 diabetes after infections (aRR 1.06, 95%CI:0.77, 1.45, Figure 2 a). There was moderate evidence for heterogeneity, with a suggestive increased risk among children in Norway only (aRR 1.80, 95%CI: 1.07, 3.02). Test-negative design analysis and results from the age-group 0-17 years in Norway was similar, with a slightly weaker and non-significant association for Norwegians aged 12-17 years using the test-negative design (aRR 1.58, 95%CI: 0.93, 2.70, Supplemental Figure S4). Unadjusted and adjusted rate ratios were largely similar (Supplemental Figure S5a), suggesting no major confounding by the measured covariates.

**Figure 2.**
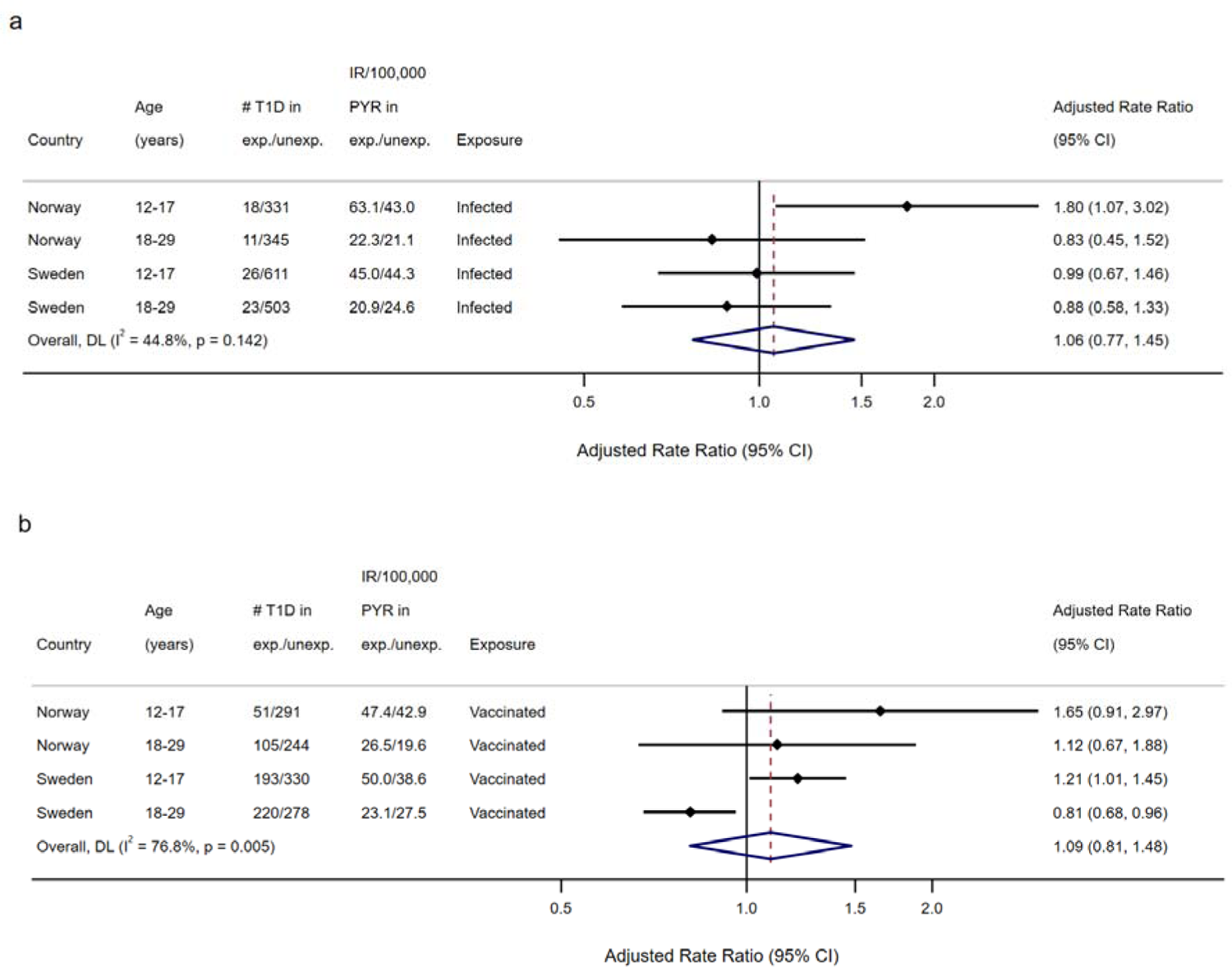
Risk of type 1 diabetes after SARS-CoV-2 infection or vaccination. **a**: Association of SARS-CoV-2 infection with risk of type 1 diabetes. **b**: Association of SARS-CoV-2 vaccination with risk of type 1 diabetes. In both a and b, rate ratios were adjusted for age, sex and country background. Additional adjustments were done for coeliac disease, family income, crowding, household size, urban/rural residence and geographical region for Norwegian data. *T1D: Type 1 diabetes; IR: Incidence Rate; PYR: Person-years. CI: Confidence interval. DL: DerSimonian and Laird random effects pooled estimate. p-values for pooled (overall) results are from Cochran’s Q-test for heterogeneity*.

In post hoc analyses among children aged 0-17 years in Norway infected with SARS-CoV-2 before 30 November 2021 (pre-omicron) the association with type 1 diabetes (aRR 1.57, 95% CI:1.00, 2.47) was similar to the association for children infected later (aRR 1.42, 95% CI: 0.76, 2.67), although the latter was not significant (Supplemental Table S3). With extended follow-up with respect to type 1 diabetes to the end of April 2024, there were no association between SARS-CoV-2 infection and type 1 diabetes among children in Norway (aRR 0.99, 95%CI: 0.87, 1.13, Supplemental Table S3).

### SARS-CoV-2 vaccination and subsequent risk of type 1 diabetes

The first vaccines were administered in December 2020, with the large majority from July 2021, somewhat later for the youngest age-group (Supplemental Figure S1). The distribution of elapsed time between SARS-CoV-2 vaccination and type 1 diabetes varied from <30 days to more than one year with no apparent clustering of cases (Supplemental Figure S2 and Figure 3b).

**Figure 3.**
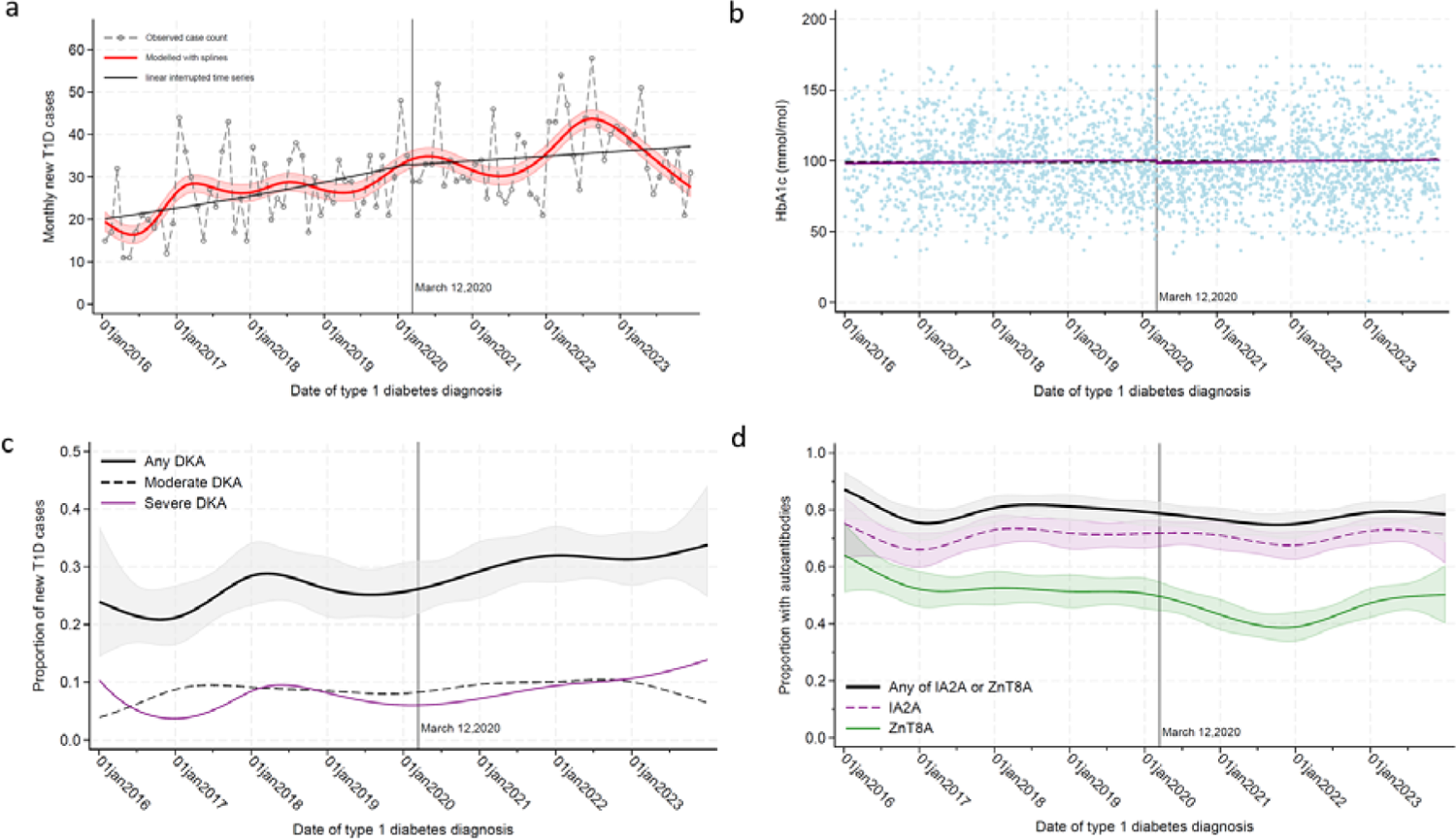
Time trend in incidence and severity of newly diagnosed type 1 diabetes 2016-2023. (T1D) in 2913 people aged below 18 years during 2016-2023 in the nationwide Norwegian Childhood Diabetes Register (NCDR). a: Monthly case counts, b: glycated haemoglobin (HbA1c) at diagnosis of T1D, c: Biochemical Diabetic Ketoacidosis (DKA) at diagnosis of T1D. Any DKA (pH < 7.3 or bicarbonate <15 mmol/L). Moderate DKA was defined as pH <7.2 or serum bicarbonate <10 mmol/L, and severe DKA was defined as pH <7.1 or serum bicarbonate <5 mmol/L. d: Autoantibodies to insulinoma antigen-2 (IA2) and Zinc Transporter-8 (ZnT8). Vertical lines represent the date of the first lockdown – 12 March 2020. Shaded areas in panel a, c and d are 95% confidence intervals for regression model predictions.

Pooled results for Norway and Sweden and age-groups 12-17 years and 18-29 years showed no significant association between SARS-CoV-2 vaccination and risk of type 1 diabetes (aRR 1.09, 95%CI: 0.81, 1.48, Figure 2b). There was significant heterogeneity, primarily driven by the opposite direction of association for children and young adults in Sweden (increased risk in children and reduced risk in young adults). Adjusted and unadjusted associations were largely similar (Supplemental Figure S5b). Additional post hoc analyses by vaccine dose showed similar results (Supplemental Figure S6).

Post hoc analyses with follow-up to the end of April 2024 in Norway showed no significant association between SARS-CoV-2 vaccination in children aged 12-17 years (aRR 1.08, 95%CI: 0.86, 1.37) or young adults aged 18-29 years (aRR 1.06, 95%CI: 0.79, 1.42, Supplemental Table S3).

### Time trends in incidence and severity of type 1 diabetes at diagnosis

Among 2913 newly diagnosed cases of type 1 diabetes under 18 years of age during 2016-2023 in Norway, the incidence increased overall throughout the study period (linear trend P<0.001, Figure 3a). There was an increase during 2022 followed by a decline to pre-pandemic rates, and a tendency towards a downward trend-break after 12 March 2020 (p=0.07, interrupted time-series). There was a significant overall seasonal variation in incidence, and a tendency towards a changing seasonal pattern after 12 March 2020 although this was not statistically significant (p=0.19 in the cosinor model).

The mean HbA1c at diagnosis was stable during 2016-2023 (Figure 3b). DKA at diagnosis of type 1 diabetes increased overall (p<0.001), but there was no significant trend break for any DKA after 12 March 2022 (p=0.93), nor for moderate or severe DKA (p>0.3, Figure 3c). This was also supported by the mean levels of the underlying pH and bicarbonate data (Supplemental Figure S7). The proportion with positive islet autoantibodies at type 1 diabetes diagnosis was stable during 2016-2023 for IA2A or any of IA2A or ZnT8A (Figure 3d). There was an overall significant downward trend for ZnT8A, and a tendency towards a trend break after 12 March 2020 (p=0.09 for interrupted time-series) although the change appeared around January 2022.

## Discussion

Using complete population health registries from two Scandinavian countries with differences in coronavirus epidemic and countermeasures, we comprehensively investigated the potential impact of the pandemic, SARS-CoV-2 infections and vaccinations on the severity at diagnosis, seasonality, and risk of developing type 1 diabetes in children and young adults.

Overall, we found no significantly increased risk of type 1 diabetes after SARS-CoV-2 infection or vaccination in the pooled analyses. There were suggestive sub-group associations of infection with increased risk type 1 diabetes in children in Norway only, and of vaccinations with increased risk of type 1 diabetes in children with opposite direction of association in young adults. While the population incidence of type 1 diabetes increased throughout the study period among children in Norway, there was no clear trend-break and no significant worsening in severity at onset of type 1 diabetes after the pandemic. Aspects of the pandemic and incidence of type 1 diabetes may be unique to each country, but we believe our overall association results are generalizable to other countries with largely European origin populations.

While it remains unclear whether SARS-CoV-2 infections or vaccinations are associated to type 1 diabetes due to biases, many potentially plausible biological mechanisms for such a link have been proposed.^10,27^ Both infections and vaccinations induce an immune response with inflammation that could potentially influence progression of ongoing islet autoimmunity. Examples include viral persistence, molecular mimicry and bystander activation of memory T-cells.^10,28^ Infections, particularly with fever, may increase insulin resistance with consequent metabolic challenge and beta-cell stress that could cause earlier presentation of diabetes and early diagnosis of at-risk individuals. The changing time trends seen in many studies may partially be explained by indirect consequences of the pandemic.^11^ Lockdown led to changes, for instance in health-seeking behaviour and exposures to other viral infections.^29^

Induction of autoimmunity by SARS-CoV-2 infections have also been suggested,^30^ although prospective evidence linking SARS-CoV-2 infections to robustly defined new onset islet autoimmunity is limited to a single study.^31^ The other two studies of islet autoimmunity likely included prevalent rather than incident autoimmunity,^32^ or too few cases of incident autoimmunity to be conclusive.^33^ Because the median time from seroconversion for islet autoantibodies to clinical type 1 diabetes is over five years in genetically susceptible children,^34^ we believe that the follow-up in our and previous cohort studies is too short for such a mechanism to explain potential associations between infections and clinical type 1 diabetes. Most infections occurred during or after the omicron variants emerged, and follow-up after this were therefore short.

### Comparison to previous studies of SARS-CoV-2 infections and risk of type 1 diabetes

Total population studies similar to ours from Denmark and Scotland found no association between SARS-CoV-2 infections and risk of type 1 diabetes,^16–18^ whereas other studies using claims databases reported 1.4 to 2-fold increased risk of type 1 diabetes.^19,35^ Details of our literature review is outlined in the Supplemental Appendix. Among ten previously published cohort studies, there was clear heterogeneity among studies (Supplemental Table S4 and Supplemental Figure S8). Our interpretation is that the body of evidence does not support a clear association between SARS-CoV-2 infection and subsequent risk of type 1 diabetes.

### Comparison to previous studies of SARS-CoV-2 vaccination and risk of type 1 diabetes

We only identified a single cohort study of SARS-CoV-2 vaccination and risk of type 1 diabetes, reporting no significant association.^16^ Other studies were case reports (Supplemental Table S5). Whether these are coincidence or causal relations is impossible to conclude. There is a need for further studies with longer follow-up after vaccination, but our interpretation of the currently available body of evidence including our data is that there is no consistent evidence supporting a relationship of appreciable magnitude.

### Comparison to previous studies of incidence and characteristics at onset of type 1 diabetes

An increase in incidence of type 1 diabetes was reported from several countries, with an overall 14% and 27% relative increase in incidence for 2020 and 2021 compared to 2019 in a meta-analysis.^11^ The time trend seems robust, but there is still a question of whether underlying time trends substantially increased also after 2021. A recent study from Scotland showed that the incidence of type 1 diabetes in children aged 6-14 increased throughout 2020, but decreased from early 2021 and returned to pre-pandemic levels by the end of 2021.^36^ Our results were largely consistent with the Scottish albeit with a later peak and subsequent decline in Norway. Longer follow-up of additional populations is needed to conclude whether similar patterns are common elsewhere. An international study from 13 diabetes registries reported an increase in the proportion with DKA in children with new onset type 1 diabetes during the COVID-19 pandemic.^15^ When we further investigated potential seasonal variation in DKA at onset of type 1 diabetes in Norway, we found no clear trend break or changes in seasonality after the first lock-down.

### Strengths and limitations

An important strength of the current study is the inclusion of two neighbouring countries with differences in pandemic countermeasures and including populations who receive free care (including testing for SARS-CoV-2) and free of charge hospitals admissions. The data came from complete population registries. The coverage of type 1 diabetes is deemed robust, as insulin is necessary for survival in this disease. However, it is unlikely that all SARS-CoV-2 infections were covered. Especially asymptomatic infections may have been missed. On the other hand, mass testing in schools occurred during part of the study period, and we ended follow-up shortly after routine testing was no longer recommended, to minimise misclassification of infections. Recommendations for laboratory testing have changed during the pandemic, with limited capacity during the first months and high capacity and highly accessible testing from August 2020 to February 2022 (see ref.^37^ and Supplemental Methods for additional details). Until mass testing was rolled out, most cases of SARS-CoV-2 in younger people were not detected, but seroprevalence studies from this period indicate very few infected.^38^

## Conclusion

No significant break in time trends after the first lockdown in March 2020 for incidence, or HbA1c, DKA diabetes ketoacidosis, and islet autoantibodies at diagnosis of type 1 diabetes was observed in Norway. While further studies and longer follow-up are warranted, our combined Norwegian and Swedish results together with other available evidence suggest no major consistent associations of SARS-CoV-2 infection or vaccinations and risk of type 1 diabetes in young people aged 12-29 years.

## Author contribution

(*CRediT format* https://credit.niso.org/)

*Conceptualization*: LCS, PLDR, GT, HLG

*Methodology*: LCS, PLDR, HLG, GT, RL, NP, AS, BZ, HB

*Formal analysis and visualization*: GT (Norwegian Beredt-C19 data), NP (Swedish data), LCS (NCDR data and meta-analyses of Swedish and Norwegian population-registry data), OG, NALB (NCDR data).

*Data Curation*: GT, OG, NALB, HB, KS, NP, RJ, AS, BZ.

Writing - Original Draft: LCS, PLDR, GT

*Writing - Review & Editing*: All authors (LCS, PLDR, RL, HB, HLG, NP, AS, BZ, KS, OG, NALB, TS, GT)

Project administration and Resources: HLG, RS, RJ

Approval of the final manuscript for submission: All authors (LCS, PLDR, RL, HB, HLG, NP, AS, BZ, KS, OG, NALB, TS, GT)

## Data Availability

Data privacy legislation in Norway and Sweden does not allow the authors to share individual level patient data. Researchers can apply for similar data to the respective registry holders after ethics approval in the respective countries. Researchers can apply for access to the Norwegian population health registry data by application to https://helsedata.no/, and for access to data from the Norwegian Childhood Diabetes by sending an application to. The Swedish Medical Products Agency will consider proposals for research collaboration. Enquiries can be submitted to the Swedish Medicines Agency ( and CC).

## Acknowledgments

We thank all those who collected and report data to the various registries and those who performed laboratory tests from which results went into the registries during the pandemic in Norway and Sweden. The authors wish to thank Nina Gunnes (previously with Norwegian Institute of Public Health) for quality control of computer code for survival analysis, especially for the time-varying covariates.

## Declaration of interests

All authors completed a conflict-of-interest form. The authors report no personal conflicts of interest pertaining to this work. Honoraria or other outside this work is described below:

Paz Lopez-Doriga Ruiz report participation in research projects funded by pharmaceutical companies (one on diabetes drug), all regulator-mandated Phase IV studies (PASS) unrelated to the submitted work, with all funds paid to their institution (no personal fees).

Dr Sundström reported participating in research funded by governmental agencies, universities, Astellas Pharma, Janssen Biotech, AstraZeneca, Pfizer, Roche, (then) Abbott Laboratories, (then) Schering-Plough, UCB Nordic, and Sobi, with all funds paid to Karolinska Institutet, outside the submitted work.

Dr Ljung reported receiving grants from Sanofi Aventis paid to his institution outside the submitted work; and receiving personal fees from Pfizer outside the submitted work.

No other disclosures were reported.

## Notes

### Author Declarations

The Norwegian Institute of Public Health is entitled by law to set up and use the preparedness register (Beredt C19) to help national and international authorities handle the pandemic. The emergency preparedness register was established according to the Health Preparedness Act paragraph 2-4. The study was approved by the Norwegian Regional Committee for Medical and Health Research Ethics South-East (REK Sor-Ost A, ref 122745), and has conformed to the principles embodied in the Declaration of Helsinki. Data from the Norwegian Childhood Diabetes Registry was routinely collected as part of the medical quality registry, and the use of data for this study together with BeredtC91 was approved by REK Sor-Ost A. Swedish Ethics approval: The Swedish study is approved by the Swedish Ethical Review Authority (2020-06859, 2021-02186) and has conformed to the principles embodied in the Declaration of Helsinki. Consent to participate is not applicable as this is a register-based study.

