## Supplemental Appendix for "Type 1 diabetes risk and severity after SARS-CoV-2 infection or vaccination"

#### Supplemental Methods

##### The Norwegian Emergency Preparedness Register for COVID-19

The Emergency Preparedness Register for COVID-19 (Beredt-C19) linkage of several total population registers. [1,2] All citizens in Norway at birth or immigration are registered with a personal identification number for administrative purposes. The personal number enables linkages of individual level information across health and administrative registries. The registry was established in 2020 to provide authorities with update information on prevalence, causal relationships, and consequences of the COVID-19 epidemic in Norway. Beredt C19 compiles updated individual-level data from several registers including information already collected in the healthcare system, national health registries and administrative registers. In this study we used data from the sources described below. Disease codes are listed in Supplemental Table S1.

*National Population Register of Norway*
The National Population Register includes information of everyone that resides or have resided in Norway (https://www.skatteetaten.no/en/person/national-registry/). We used date of birth, immigration, emigration, and death from this register.

*Statistics Norway: individual level sociodemographic data*
Statistics Norway (https://www.ssb.no/en) keeps records of sociodemographic variables including level of education, country of birth, country background (country of birth of parents and grandparents), and family number used for robust variance estimates accounting for intra-familial correlations.

*The Norwegian Patient Registry (NPR)*
The NPR includes individual level information on all contacts with non-primary healthcare services. Information registered includes admission and discharge dates, and diagnostic codes during the hospital stay and outpatient contact. These codes are according to the International Classification of Diseases version 10 (ICD-10).[3] We used E10 code (type 1 diabetes) from inpatient or outpatient stays or contacts with private-practicing specialists. E11-E14 (other diabetes types) were used to exclude people with diabetes before study start or to censor people who developed non-type 1 diabetes during follow-up.

*Control and Payment of Health Reimbursement (KUHR) database for primary care in Norway*
The KUHR database covers health services financed by the state, bills are the basis for the information stored in KUHR (https://helsedata.no/en/forvaltere/norwegian-directorate-of-health/norwegian-control-and-payment-of-health-reimbursements-database-kuhr/). We retrieved diagnoses (based on ICPC-2 codes) and diagnosis dates from this database. T89 code was used for type 1 diabetes, and T90 (type 2 diabetes) was used to exclude people with diabetes before study start or to censor people who developed non-type 1 diabetes during follow-up.

*Norwegian Prescription Database (NorPD)*
All prescribed medicines in Norway dispensed from a pharmacy are registered in the NorPD. We retrieved the date of collection and the product classified using the Anatomical Therapeutic Chemical code (ATC codes) A10A for insulins, whereas use of A10B (non-insulin glucose lowering medication) was used to exclude other types of diabetes (Supplemental Table S1).[4]

*Norwegian Surveillance System for Communicable Diseases (MSIS) and the MSIS laboratory database*
COVID-19 is a mandatory notifiable disease and all cases in Norway should be reported to MSIS. In addition, reporting of both positive and negative SARS-CoV-2 tests are mandatory to the MSIS laboratory database which contains both date of testing and test results. We retrieved dates of all registered laboratory-confirmed cases of SARS-CoV-2 infections. Most of the test results were based on polymerase chain reaction (PCR), a small proportion were antigen tests. Recommendations and capacity for testing changed during the study period, from being restricted in the first weeks of the pandemic to widely accessible from August 2020. Mass testing in schools occurred during part of the study period, but the general recommendation to test for SARS-CoV-2 was removed by end of January 2022 and testing frequency gradually declined over the subsequent two months. Subsequent reporting has been largely from hospitalized patients. All testing was free of charge and easily accessible to ensure a high uptake.[5,6]

*The Norwegian Immunisation Register (SYSVAK)*
SYSVAK is a register of vaccines in Norwegian vaccination programs, with mandatory registrations of all COVID-19 vaccinations https://www.fhi.no/en/va/norwegian-immunisation-registry-sysvak/. The information includes personal identifier and date of vaccination. The vaccine products are specified by a unique identifier

based on a combination of brand name, substance, formulation.[7] Detailed descriptive data on number of vaccines by age-group, dose and type of vaccine can be retrieved from the NIPH statistics website (in Norwegian): https://allvis.fhi.no/sysvak. We used SYSVAK-data to identify recipients of the BNT162b2 (Pfizer-BioNTech) or mRNA-1273 (Moderna) vaccine. Vaccines against COVID-19 were offered to individuals aged 12 years or older. Use of other vaccines in the included age-groups was very rare (<1000 vaccinations) except the AstraZeneca vaccine which was given to around 50,000 young adults during 2021 before it was suspended in Norway https://allvis.fhi.no/sysvak .[8] Norway recommended and offered those aged 12-15 years a single dose of BNT162b2 from 2 September 2021, and a second dose from the end of January 2022,[5] while only around 16% of individuals aged 12-15 who received the first dose also took a second dose https://allvis.fhi.no/sysvak. Those aged 16-17 years were recommended and offered two doses of BNT162b2 from August 18, 2021.[5] Around 95% of those aged 16-17 years who took a first dose also took a second dose https://allvis.fhi.no/sysvak.

#### Swedish COvid-19 VACcination register SAFEty study in Sweden

Individual level linkage of several health registries were combined for analysis in the Swedish COvid-19 VACcination register SAFEty study in Sweden (CoVacSafe-SE), as described in detail elsewhere.[9] We used data from Swedish residents aged 12-29 years in 2020, for the period 2015 to 2022, from registries described below as in ref.[9]

*The Total Population Register*

The Total Population Register contains information on personal identifier, date of birth, country of birth, place of residence, marital status, date of death, and date of immigration and emigration. The register is held by Statistics Sweden.

*The national vaccination register of Sweden*

The national vaccination register contains since 1 January 2021 information on vaccination for COVID-19. The information includes personal identifier and date of vaccination. The vaccine products are specified by a unique identifier based on a combination of brand name, substance, formulation, batch number, and dose number (for repeated doses). The completeness of registration within the child immunization program is high, where 98.4% of children had at least one vaccination recorded, however, completeness of registration of vaccination for COVID-19 has not been published. The register is held by the Public Health Agency of Sweden. Vaccination register and vaccination coverage - The Public Health Agency of Sweden (folkhalsomyndigheten.se)

*Register on surveillance of notifiable communicable diseases (SmiNet)*

SmiNet contains information on notifiable diseases which must be reported by the laboratories and the physician treating the patient, or performing an autopsy, in accordance with the Swedish Communicable Diseases Act18. SmiNet includes personal identifier, date of disease occurrence, date of testing, date of positive test, and diagnosis of notifiable infectious disease. The register is held by the Public Health Agency of Sweden.

*The Swedish national patient register*

The Swedish patient register comprises information on all in-hospital care and out-patient specialist care in Sweden. The information includes personal identifier, admission and discharge dates, whether hospitalisation was planned or acute, codes for discharge diagnoses and surgical procedures, whether discharged as deceased, to own private residence or other health care facilities, type of department, and hospital. It has nation-wide coverage regarding in-patient care since 1987 and specialized outpatient care since 2001. The in-hospital part of the register is a discharge register; hence, the admission is reported at time of discharge. There is no information on admission diagnoses. There is no information on primary care. During the study period diagnoses were recorded according to the Swedish clinical modification of the 10th revision of the International Statistical Classification of Diseases and Related Health Problems (ICD-10-SE). There are monthly updates, but there is a lag in reporting to the register, for the last four weeks before 3 October 2021, the completeness compared to previous years was 91%, 81%, 75%, and 58% respectively. We have no reason to believe that reporting from health care providers and county councils to the National Board of Health and Welfare are dependent on either vaccine status or outcome. The register is held by the National Board of Health and Welfare.

*The Swedish Prescribed Drug Register*

The Swedish Prescribed Drug Register contains details of all the prescriptions dispensed in Sweden since July 1, 2005. It is updated monthly with around 100 million prescriptions dispensed each year. It covers the entire Swedish population and includes information on unique personal identifier of the patient, age, sex, place of residence, and prescription information on substance, brand name, formulation and package dispensed amount, dosage (in free text) and unique expenditure and reimbursement, date of prescribing and dispensing, practice that has issued the prescription, and prescriber's profession. Drugs are identified by a unique identifier for each specific combination of brand name, substance, formulation, and package. Additionally, all drugs are classified according to the Anatomic Therapeutic Chemical Classification System (ATC). The register only includes filled prescriptions, not medicines sold over the counter, nor medicines administered directly by health-care personnel without prescription. The register is held by the National Board of Health and Welfare.

##### Outcome definition in the total population registers: New-onset type 1 diabetes

For the total population cohorts in Norway and Sweden, the endpoint was diagnosis of incident type 1 diabetes. This was defined as a type 1 diabetes diagnosis code (ICD-10 code E10 or ICPC-2 code T89) plus use of insulin (ATC-code A10A, see Supplemental Table S2 for list of codes). According to clinical practice in Norway and Sweden, we required an initial hospital stay for type 1 diabetes diagnosed before 18 years of age, but not for diagnosis at age 18-29 years. For young adults, but not children, we allowed use of non-insulin glucose lowering drugs (ATC code A10B) in addition to insulins (A10A). Any other diabetes codes (ICD-10 E11-14, ICPC-2 T90) were excluded. The earliest type 1 diabetes disease code or dispensed insulin prescription was used as the date of diagnosis (for those diagnosed after 2022 in post hoc robustness analyses, we did not have access to medication data but in practice the first type 1 diabetes disease code was the appropriate for defining date of diagnosis). To avoid including prevalent cases, we excluded individuals with a previous diabetes diagnosis or previous use of any glucose-lowering medications during 2017-2019 in Norway and 2015-2019 in Sweden.

##### The Norwegian Childhood Diabetes Registry

The Childhood Diabetes Registry (NCDR) is a population-based registry including all incident cases of diabetes diagnosed before age 18 years, and monitors quality indicators of diabetes care. Signed consent is obtained from each patient and/or the parents (less than 1% refuses to be registered). Classification into type 1 diabetes, type 2 diabetes, monogenic or other types (including syndromic and secondary diabetes) is done based on a combination of degree of glycaemia (glycated haemoglobin, HbA1c and non-fasting glucose), non-fasting C-peptide), islet autoantibodies, and suspected cases of monogenic diabetes is diagnosed based on family history and genotyping. Over 90% of newly diagnosed diabetes are clinically judged to be type 1 diabetes and included in the current analyses, and ascertainment has been documented to be above 95% by comparison to both the Norwegian Patient Registry and Norwegian Prescription Database, described in more detail below. Date of diagnosis of type 1 diabetes is defined as the first day of insulin treatment, in accordance with EURODIAB criteria.[10-13]

*Laboratory measurements at diagnosis of type 1 diabetes*

Blood pH, serum bicarbonate concentration ($HCO_3^-$) and HbA1c was measured at the clinical chemistry department in the diagnosing hospitals using standard methodology (e.g. Cobas multianalyte instrument from Roche or High Pressure Liquid Chromatography, HPLC). Blood pH and serum bicarbonate were used to define Diabetic Ketoacidosis (DKA), including subdivided in to mild, moderate and severe, according to the International Society for Pediatric and Adolescent Diabetes (ISPAD) 2018 criteria.[14] Updated ISPAD criteria were published in 2022,[15] but was not used here since these did not apply to most of the study period included in this paper. The ISPAD 2018 criteria for biochemical DKA (in addition to hyperglycaemia present at diagnosis of type 1 diabetes) were venous blood pH <7.3 or bicarbonate <15 mmol/L (ketonemia or ketonuria or clinical symptoms of DKA were not considered in the current analysis). Furthermore, criteria for mild DKA were venous pH <7.3 or serum bicarbonate <15 mmol/L, moderate: pH <7.2 or serum bicarbonate <10 mmol/L, and severe: pH <7.1 or serum bicarbonate <5 mmol/L.[14]

Data on time trends of DKA at diagnosis of type 1 diabetes in NCDR by calendar year before and after start of the corona pandemic has previously been published as part of a multi-centre study.[13] However, in the current paper, we analysed these data (together with underlying trends in pH, bicarbonate and severity of DKA, and other biomarkers at diagnosis) at a higher resolution by calendar month of diagnosis to account for known underlying seasonal variations in some of these, and possibly changes in the seasonality during the pandemic.

Islet autoantibodies in serum taken at diagnosis was measured in duplicate centrally at the Hormone laboratory at Oslo University hospital. During the study period, autoantibodies against four islet autoantigens were measured – the intracellular domain of the receptor type protein phosphatase N usually called IA2, zinc transporter 8 (ZnT8), the 65kD glutamic acid decarboxylase 2 (GAD2), and insulin.[16] However, because the Hormone laboratory changed the assays used for autoantibodies to GAD2 and insulin in 2020, consistent time trends for islet autoantibodies could only be studied for autoantibodies to ZnT8 and IA2 in the current analyses. These were measured using in house radio-binding assays as described in detail elsewhere.[17,18] The hormone laboratory participates regularly in the Islet Autoantibody Standardization Programme, an external quality control programme with ring testing of pooled sera from patients with type 1 diabetes and controls.[19] Among a total of 2913 newly diagnosed individuals during 2016-2023, 66 (2.3%) had missing data for pH and bicarbonate, 96 (3.3%) had missing HbA1c, 108 (3.7%) had missing IA2A and 142 had missing ZnT8A (4.9%).

#### Ethics

The Norwegian Institute of Public Health is entitled by law to set up and use the preparedness register (Beredt C19) to help national and international authorities handle the pandemic. The emergency preparedness register was established according to the Health Preparedness Act §2-4. The study was approved by the Norwegian Regional Committee for Medical and Health Research Ethics South-East (REK Sør-Øst A, ref 122745), and has conformed to the principles embodied in the Declaration of Helsinki. Data from the Norwegian Childhood Diabetes Registry was routinely collected as part of the medical quality registry, and the use of data for this study together with BeredtC91 was approved by REK Sør-Øst A. Swedish Ethics approval: The Swedish study is approved by the Swedish Ethical Review Authority (2020-06859, 2021-02186) and has conformed to the principles embodied in the Declaration of Helsinki. Consent to participate is not applicable as this is a register-based study.

#### Supplemental statistical analysis

*Analysis of association between SARS-CoV-2 infection or vaccination and risk of type 1 diabetes*

The initial analysis was planned for and done with Norwegian data only, while the Swedish data were subsequently included for replication and overall increased robustness of results. This approach influenced the detailed analysis plan. For instance, the Swedish group had previously used Poisson regression, whereas the Norwegian group had used Cox regression, although the two methods usually produce equivalent results as long as the time variable is flexibly modelled in Poisson regression.[20] The a priori analysis plan for the Norwegian SARS-CoV-2 infection data specified test-negative design as the primary analysis to account for potential selective testing occurring early in the pandemic, but because most of the population were later tested and Norwegian analyses showed little difference between test-negative design results and full cohort results (see main Figure 3a), Swedish data were only analysed using full cohort design. Individuals were followed from entering a given age-category to diagnosis of type 1 diabetes, emigration, death or end of follow-up (28 April 2022 in the primary analyses and later in extended robustness analyses), whichever occurred first.

The Swedish full population linked registry cohort included only individuals aged 12 or above, and we could thus not assess the potential association between SARS-CoV-2 infection and type 1 diabetes in children below 12 in Sweden. This should have little impact on the result for vaccines because only Norwegian children aged 12 or above were vaccinated in Norway (and Sweden). Based on considerations of expected number of newly diagnosed cases of type 1 diabetes and frequency of infections and vaccinations in the study period, we decided a priori not to stratify association analyses by sex or age-groups subdivided in finer categories than 0-17 and 18-29. However, because Swedish data were not available for individuals younger than 12 years, we therefore estimated associations in the Norwegian data also for comparable age-groups to be able to directly compare with Swedish results. In both countries children received mRNA vaccines against SARS-CoV-2. However, while Norwegian children aged 12-17 mostly got a single dose of SARS-CoV-2 vaccine, a large proportion of 12-17-year-olds in Sweden received two doses. We therefore analysed associations per first and second dose as a robustness analysis. In both countries, we knew that hospitalizations for COVID-19 among children were too rare to be meaningfully analysed for association with subsequent risk of type 1 diabetes.

We adjusted all models for sex, age and Nordic vs non-Nordic country of origin – variables we consider plausible confounders potentially associated with both exposure and outcome. We also mutually adjusted for SARS-CoV-2 infection and vaccination in all models. We also adjusted for coeliac disease diagnosis at study start, low family income (EU60, below 60% of the median household income adjusted for household composition), crowded housing, household size, urban/rural residence and regions (South-East, West, Mid, and North) in Norway. These variables are primarily believed to predict risk of infections, and are not established risk factors for type 1 diabetes, but were included to assess robustness of results towards adjustment for these variables. (There were relatively small differences between adjusted and unadjusted associations of both SARS-CoV-2 infections and vaccinations with type 1 diabetes in both countries (see Supplemental Figure S4 below), suggesting that this should have limited impact). Analyses of Norwegian data were done using Stata (StataCorp. 2021. Stata Statistical Software: Release 17. College Station, TX: StataCorp LLC) and Swedish data were analysed using SAS version 9.4 (SAS Institute Inc). All analyses were complete case (there was generally a low proportion with missing data on covariates, see Supplemental Table S2).

*NCDR data on incidence and clinical characteristics of newly diagnosed type 1 diabetes*
Analysis of NCDR data on incidence and severity at onset of type 1 diabetes by date of diagnosis before age 18 years were analysed by plotting observed monthly counts, modelling with splines, and interrupted time-series. All analyses were complete case (no imputation of missing data). Monthly case counts were modelled with negative binomial regression, continuous variables pH, bicarbonate and HbAc1 were modelled using linear regression models, whereas the dichotomous measures DKA and islet autoantibody positivity were modelled using logistic regression models. Time trends were modelled with restricted cubic splines with knots every six months except in the beginning and end of the study period (31 January 2016, 1 July 2016, 1 January 2017, 1 July 2017, 1 January 2018, ……, 1 July 2022, 1 January 2023, 1 July 2023, 1 December 2023). To assess robustness for the time trend estimation using restricted splines with this selection of knots, we also used plotted similar predictions using splines with automated selection of smoothness parameters as implemented in the R package *mgcv*: Mixed GAM Computation Vehicle with Automatic Smoothness Estimation (https://CRAN.R-project.org/package=mgcv) .[21] This produced very similar results. Seasonal variation in case-counts and clinical characteristics at diagnosis of type 1 diabetes were assessed using cosinor models with sine- and cosine terms for functions of day (1-366) of year.[22] Potential changes in the seasonal pattern after onset of the first lockdown 12 March 2020 was assessed by interaction terms between the sine and cosine terms and an indicator of before/after this date. Two degrees of freedom likelihood ratio tests comparing the model with and without interaction terms were used as a formal test of changing seasonal pattern. Potential interruptions in underlying time trends for incidence and clinical characteristics at diagnosis of type 1 diabetes were assessed using interrupted time trend analysis.[23] Based on previous experience with uncertainty (random variation) around incidence estimates, especially when investigating seasonal effects, we decided a priori not to stratify data by gender and age-groups within the available 0-17 year age-group.

#### Literature review and quality of previous evidence

This section provides a detailed outline of the search strategy, publications identified and evaluation of the quality of published evidence linking SARS-CoV-2 infection or vaccination to subsequent risk of type 1 diabetes. An early feature of COVID-19 research was the mass production of systematic reviews of this condition, with various degree of focus and generally poor reporting. By April 2020 there were 513 registered systematic review protocols in the international Prospective Register of Systematic Reviews many with severe limitations.[24] In the fields of new-onset type 1 diabetes after SARS-CoV-2 infection, we identified two systematic reviews published in 2021 [25,26] and six published in 2022,[27-31]Lai, 2022 #754} and three published systematic reviews in 2023.[32-34] Fewer studies have investigated the risk of type 1 diabetes after SARS-CoV-2 vaccinations, and the majority of original publications are case-reports.[35,36] Many published systematic reviews were not making distinctions between important scientific concepts outlined below, and the number of high quality-original cohort studies in this field is limited.

*Types of study and quality of evidence*
Hierarchy of study designs: Randomised trials represent the gold standard for quality of evidence for causal effects of interventions. Because the primary vaccine efficacy trials did not include type 1 diabetes as outcomes and infections cannot be studied experimentally in humans with respect to risk of type 1 diabetes, the types of studies are limited to observational human studies, or experimental studies in animal models or in vitro studies. The latter provide indirect evidence (briefly reviewed in the introduction of the main paper). We evaluate here human observational studies. A traditional hierarchy of evidence for human observational studies ranges case-reports lowest, case-control studies intermediate and prospective cohort studies highest. Another type of study are descriptive epidemiological or clinical studies of time trends in type 1 diabetes incidence before and after the start of the pandemic. The latter type of study can be informative even if no individual data are available on exposure to SARS-CoV-2 infection or vaccination. They should ideally include standardized methods for ascertaining new onset type 1 diabetes in well-defined geographical catchment areas or populations and a resolution in calendar time that allows investigation of seasonal variations. This is because seasonal variation normally occurs, and this could potentially have been different after the start of the pandemic.

Bias in observational studies: The major forms of bias are selection bias, measurement error and confounding. While both randomized trials and observational studies can suffer from selection bias and measurement error in outcome, observational studies are especially vulnerable to confounding. Potential confounding cannot be eliminated in observational studies, except under strong unverifiable assumptions. Prospective cohort studies controlling potential confounding factors can minimize the risk of bias including selection bias. Confounding variables are variables that are causally associated with both exposure (SARS-CoV-2 infection or vaccination) and outcome (risk of new onset type 1 diabetes). Typical factors that may be confounders are age and ethnicity/immigration status. While family history of type 1 diabetes is associated with risk of disease, it is typically relatively rare and not necessarily associated with risk of infection or vaccination so are likely to be a weak confounder, if at all. Measurement error can occur in both exposure, covariates and outcome variables. While type 1 diabetes in young adults are usually considered reasonably easy to ascertain, it is important, especially in registry-based studies, to make sure that registered type 1 diabetes diagnosis is actually new onset because ICD-10 and other disease codes do not differentiate between incident and prevalent cases. A look-back period of at least 1-2 years is typically required for children, and possibly more for adults to ensure that a registered type 1 diabetes disease code represent a new onset case. A few studied used islet autoimmunity as the outcome, and it is important to define this robustly with confirmed multiple islet autoantibodies verified to be negative at the time of exposure.

Exposure measurement: While SARS-CoV-2 vaccinations represent well-defined events that is easy to remember and is often registered in databases, these will typically not suffer from severe measurement error. However, measurement of SARS-CoV-2 infections is not trivial. Infections typically vary in severity from asymptomatic to life-threatening, and the extent to which infections are registered in databases typically depend on the severity. Many countries had large scale screening with nasopharyngeal PCR and later antigen tests, and test results were registered in databases in many countries. Asymptomatic infections could be detected with screening in schools or defined risk groups in the population. Antibody tests could detect prior infections and/or vaccination. All of the abovementioned types of exposure assessments were considered acceptable in our literature review.

Induction or latency period: It is well established from natural history studies that the disease process in type 1 diabetes is long, with islet autoimmunity starting postnatally and a median time from islet autoimmunity to clinical type 1 diabetes of at least 5-7 years in individuals with multiple islet autoantibodies.[37] Therefore, type 1 diabetes occurring within a month or earlier after exposure (to SARS-CoV-2 infection or vaccination) is unlikely to represent a true causal effect of an aetiological factor. The required latency period is impossible to know with certainty, but most studies in the field has used outcomes after 30 days as to define the post-acute phase. Longer periods are probably even better for type 1 diabetes, but problems with longer periods include i. you can miss some true aetiological cases and ii. few studies have had the chance to have a very long follow-up period of sufficiently large numbers of individuals to allow for reasonable statistical power. An observed association between exposure to SARS-CoV-2 infection or vaccination may be due to precipitation of subclinical disease in a small fraction of the population who would otherwise have been diagnosed soon, or an increase in the rate of progression for islet autoimmunity in normoglycaemic people with multiple islet autoantibodies. Although not very likely, exposures could also slightly raise the risk of type 1 diabetes in the general population homogenously across individuals regardless of underlying risk or stage of disease.

Sample size and statistical power: The statistical power to detect an association between exposure and risk of type 1 diabetes critically depends on prevalence of exposure and number of individuals developing type 1 diabetes during follow-up. The incidence is relative rare, for instance around 30-60 per 100,000 person-years among children in the Nordic countries who have the highest incidence, and around 3-10 per 100,000 in many countries in Asia. This means that for a one-year observation period of individuals initially free of type 1 diabetes, the proportion expected to develop type 1 diabetes would be maximum 0.06% (60 new cases in a cohort of 100 000), and in most instances much smaller. If only a fraction of the cohort is exposed, for instance 50%, the expected number of exposed cases under the null hypothesis will be 30 in such a cohort. This illustrates that very large samples of population-based studies are required for reasonable statistical power. For instance, such a cohort in a high-risk population of N=100,000 of which with 50% exposed individuals would be able to detect relative risks of approximately 2.3 or higher, with a set statistical power of 80%.

*Search strategy*

Several of the authors followed publications in the field non-systematically during the pandemic, by a combination of PubMed searches, participation at scientific conferences and social media including X/Twitter and saved the publications in EndNote Databases. Authors LCS, PLDR and GT initially harmonized their relevant publications and critically reviewed the publications for relevant content. In addition, we formally search the literature from April 2020 to December 2022 using PubMed (https://pubmed.ncbi.nlm.nih.gov) with the following search terms: "diabetes[Title] AND ( COVID[Title] OR SARS-CoV-2[Title] OR pandemic[Title] OR corona[Title])". We did additional searches replacing "diabetes" with "autoimmune" or "autoimmunity", and with additional requirement of "vaccine OR vaccination", as well as separate searches for systematic reviews of the above topics. We initially screened titles for relevance. We restricted our interest to studies focusing on human type 1 diabetes in young individuals as the outcome (primarily below 30 years of age, but also including studies with older age if sub-group analyses overlapping the age-group 0-30 years were shown). We focused on prospective cohort studies (or prospective nested case-control studies within established cohorts). The search was updated for new publications up to 30 May 2024, and a final check for updates 20 June 2024. From identified original publications and reviews, we searched the reference lists for potential additional publications. In the final search we identified a high quality narrative review that included most of the original publications we had identified.[38]

*Summary of previous studies of time-trends in type 1 diabetes incidence or severity at onset*
Several papers have been published, summarized by D'Souza et al 2023.[39] They concluded from 42 studies totalling over 100,000 incident cases of type 1 diabetes among children aged below 18 years that "incidence rates of type 1 diabetes and DKA at diabetes onset in children and adolescents were higher after the start of the COVID-19 pandemic". Incidence rates has increased over many years across the globe, with year-to-year variation as well as seasonal variation. Most studies compared yearly incidences before and after onset of the pandemic, including a recent study from Finland,[40] but few investigated potential changes in seasonal variation in incidence.[41] Similarly for Diabetic Ketoacidosis (DKA) and other indices of severity of onset, most studies compared yearly levels before and after the onset of the pandemic and not potential changes in seasonal variation potentially induced by the virus, vaccinations or indirect effects of the pandemic.[13]

##### Summary of previous cohort studies of SARS-CoV-2 **infections** and type 1 diabetes risk

Prospective cohort studies of associations between SARS-CoV-2 infections and risk of type 1 diabetes are listed in Supplemental Table S4. Three identified cohort studies showed results for all ages or only adults without presenting sub-group analyses close to <30 years were not shown in the table, Syed 2023[42] Tesch 2023 [43] and Xie, 2022,[44] all of which were based on electronic health records and showed significant relative risks of 1.25-1.6. All had high incidence rates suggesting potential prevalent or false positive cases of type 1 diabetes. The Norwegian data on infections and type 1 diabetes in children aged 0-17 were presented at the EASD-conference September 2022.[45] At that time, Scottish registry data had been published (McKeigue 2022 [46]). Danish data were first published by Noorzae 2022 for age <18 years,[47] and later by Zareini 2023 who included age-groups below 30 years,[48] and both studies showed similar results (no association between SARS-CoV-2 infection and risk of type 1 diabetes). The US claims databases HealthVerity was first used in a study by Barret et al published in January 2022 not distinguishing between types of diabetes and presenting data suggesting misclassification of diabetes because of too high numbers of incident cases (not shown in the Table), and later in a similar analysis restricted to type 1 diabetes by Kompaniyets in August 2022 [49] shown in the Table (Supplemental Table S4). A registry based cohort study from Hong Kong included too few cases with type 1 diabetes to be informative and were not listed in the table.[50] The TriNetX claims database has been used in three different publications on the same topic, first Pietropaolo et al in March 2022,[51] by Kendall 2022 [52] and by Chang 2023.[53] The Cerner claims database have been used in two publications, Qeadan 2022 [54] and Bull-Otterson 2022,[55] respectively. Among the ten publications based on cohort studies listed in the table, three adjusted or stratified their analyses for SARS-CoV-2 vaccination, one excluded individuals who were vaccinated during the study period[53], while six did not include data on SARS-CoV-2 vaccination: Kompaniyets,[49] Pietropaolo,[51] Kendall,[52] Qeadan,[54] Weiss,[56] and Bull-Otterson.[55]

In summary, there were seven previous cohort studies with data for children showing relative risk point estimates ranging from 0.85 to 2.04, of which four showed a significant positive association. Two publications from the same data source, Pietropaolo and Kendall gave very different results for the same age-group. Among young adults, there were seven publications of which there were two publications each from two databases with some differences within database. Point estimates of relative risks ranged from 0.69 to 2.80 and three publications showed significant positive associations. None of the studies showing the significant positive associations with infections in at least one of the age-groups had adjusted for SARS-CoV-2 vaccinations[49,51,52,54-56]. There was substantial heterogeneity between studies overall both in methodology and associations, and a tendency that publications based on claims databases produced the strongest positive estimates that were also apparently precise (narrow 95% confidence intervals). **Although there were more positive than inverse associations, we interpret the body of evidence from previous prospective studies to be conflicting for both children and young adults with no clear overall support for the hypothesis that SARS-CoV-2 infections are associated with risk of type 1 diabetes.**

##### Summary of previous cohort studies of SARS-CoV-2 **vaccinations** and type 1 diabetes risk

As mentioned in the previous paragraph, only few of the previously published papers of SARS-CoV-2 infections had adjusted for SARS-CoV-2 vaccinations, and among them only a single study showed results for the potential association between vaccination and risk of type 1 diabetes, namely McKeigue et al. using population-based registry cohort data from Scotland. The hazard ratio comparison of the type 1 diabetes rate among those how had received one or more vaccine dose versus none, primarily based on young adults because there were few vaccinated children in the cohort, was 0.93, 95% CI: 0.73, 1.18).[57] Chang et al.[53] (using TriNetX claims database) excluded individuals who received a SARS-CoV-2 vaccine during follow-up “for clarity of interpretation” of the association analyses of infections with risk of type 1 diabetes, and the reason for this was because “the association between COVID-19 vaccination and subsequent autoimmune phenomena is controversial”, citing a review on the topic,[58] which itself cited a paper with a single case-report. This seems to be typical for the literature in this field – it is mostly based on case-reports which represent low quality of evidence in a setting where millions have been vaccinated. In most high-income countries the majority of the population has been vaccinated and anyone developing type 1 diabetes after 2021 or 2022 are highly likely to have received at least one SARS-CoV-2 vaccine. Supplemental Table 5 lists eight published papers identified in our early searches, each reporting on one or a few cases of type 1 diabetes occurring after SARS-CoV-2. Another report with two cases was published in 2024,[59] and there are probably many more.

Of two recent systematic reviews,  He et al[35] reported mainly case reports suggesting that SARS-CoV-2 could worsen glucose control in people with existing diabetes but could not conclude anything about a potential risk of new onset type 1 diabetes, while Alsudais identified five publications describing 12 patients who developed type 1 diabetes after SARS-CoV-2 vaccination but obviously could not conclude with respect to a causal relationship.[36] A registry based cohort study from Hong Kong included too few cases with type 1 diabetes to be informative and were not listed in the table.[50]

**We conclude that there is a lack of studies of the potential link between SARS-CoV-2 vaccination and subsequent risk of type 1 diabetes.**

#### Supplemental Tables

Supplemental Table S1. Disease and medication codes

| **Type 1 diabetes** | | |
|---|---|---|
| **Norwegian Patient Registry and Swedish National Patient register** | **The Norwegian Prescription Database and Swedish prescribed drug register** | **Primary healthcare database (Norway)** |
| ICD-10 code E10 | Medication ATC codes:<br>A10A - insulins | ICPC-2 codes T89 |
| **SARS-CoV2 diagnosis and vaccination** | | |
| **Surveillance System for Communicable Diseases (MSIS), MSIS-laboratory database, and Register on surveillance of notifiable communicable diseases (SmiNet)** | **Norwegian Immunisation Register (SYSVAK) and National vaccination register of Sweden** | |
| Date and result of laboratory test for SARS-CoV2 | Dates and products name of vaccines | |

ICD-10 codes: International Classification of Diseases, version 10. ACT: Anatomical Therapeutic Chemical Classification. ICPC-2: International Classification of Primary Care, Second edition.

Supplemental Table S2. Characteristics of the total population registry cohorts

| **Norway** | Age-group | | | |
|---|---|---|---|---|
| | 0-11 years | 12-17 years | 18-29 years | Total (0-29 years) |
| N | 745,405 (37.5%) | 376,665 (19.0%) | 864,900 (43.5%) | 1,986,970 (100.0%) |
| Sex | | | | |
| Female | 362,874 (48.7%) | 183,609 (48.7%) | 417,978 (48.3%) | 964,461 (48.5%) |
| Male | 382,531 (51.3%) | 193,056 (51.3%) | 446,922 (51.7%) | 1,022,509 (51.5%) |
| Country background* | | | | |
| Nordic | 446,403 (59.9%) | 260,311 (69.1%) | 632,995 (73.2%) | 1,339,709 (67.4%) |
| Europe (excl. Nordic) | 108,809 (14.6%) | 46,010 (12.2%) | 89,756 (10.4%) | 244,575 (12.3%) |
| Asia | 64,750 (8.7%) | 30,282 (8.0%) | 61,648 (7.1%) | 156,680 (7.9%) |
| Sub-Saharan Africa | 34,755 (4.7%) | 13,960 (3.7%) | 29,083 (3.4%) | 77,798 (3.9%) |
| M. East and N. Africa | 19,424 (2.6%) | 9,493 (2.5%) | 18,187 (2.1%) | 47,104 (2.4%) |
| N. Am. and Oceania | 18,163 (2.4%) | 8,399 (2.2%) | 18,144 (2.1%) | 44,706 (2.2%) |
| Latin-America | 13,862 (1.9%) | 6,090 (1.6%) | 10,844 (1.3%) | 30,796 (1.5%) |
| Missing | 39,239 (5.3%) | 2,120 (0.6%) | 4,243 (0.5%) | 45,602 (2.3%) |
| Crowded dwelling† | | | | |
| Yes | 138,622 (18.6%) | 61,269 (16.3%) | 113,051 (13.1%) | 312,942 (15.7%) |
| No | 553,882 (74.3%) | 306,694 (81.4%) | 726,973 (84.1%) | 1,587,549 (79.9%) |
| Unspecified | 12,124 (1.6%) | 5,997 (1.6%) | 18,622 (2.2%) | 36,743 (1.8%) |
| Missing | 40,777 (5.5%) | 2,705 (0.7%) | 6,254 (0.7%) | 49,736 (2.5%) |
| Low-income household‡ | | | | |
| No | 613,705 (82.3%) | 332,401 (88.2%) | 661,207 (76.4%) | 1,607,313 (80.9%) |
| Yes | 90,923 (12.2%) | 41,559 (11.0%) | 197,439 (22.8%) | 329,921 (16.6%) |
| Missing | 40,777 (5.5%) | 2,705 (0.7%) | 6,254 (0.7%) | 49,736 (2.5%) |
| Region of residence | | | | |
| Mid | 98,569 (13.2%) | 50,359 (13.4%) | 119,071 (13.8%) | 267,999 (13.5%) |
| North | 60,229 (8.1%) | 32,206 (8.6%) | 76,817 (8.9%) | 169,252 (8.5%) |
| South-East | 416,259 (55.8%) | 210,104 (55.8%) | 480,714 (55.6%) | 1,107,077 (55.7%) |
| West | 165,815 (22.2%) | 81,947 (21.8%) | 181,112 (20.9%) | 428,874 (21.6%) |
| Missing | 4,533 (0.6%) | 2,049 (0.5%) | 7,186 (0.8%) | 13,768 (0.7%) |
| Municipality size | | | | |
| < 50,000 inhabitants | 394,797 (53.0%) | 206,828 (54.9%) | 412,314 (47.7%) | 1,013,939 (51.0%) |
| ≥ 50,000 inhabitants | 346,075 (46.4%) | 167,788 (44.5%) | 445,400 (51.5%) | 959,263 (48.3%) |
| Missing | 4,533 (0.6%) | 2,049 (0.5%) | 7,186 (0.8%) | 13,768 (0.7%) |
| Coeliac disease | | | | |
| No | 743,367 (99.7%) | 374,782 (99.5%) | 863,651 (99.9%) | 1,981,800 (99.7%) |
| Yes | 2,038 (0.3%) | 1,883 (0.5%) | 1,249 (0.1%) | 5,170 (0.3%) |
| **Sweden** | | Age-group | | |
| | 0-11 years | 12-17 years | 18-29 years | Total (12-29 years) |
| N | n.a. | 703,816 (33.5%) | 1,396,372 (66.5%) | 2,100,188 (100%) |
| Sex | | | | |
| Female | n.a. | 341,647 (48.5%) | 668,749 (47.9%) | 1,010,396 (48.1%) |
| Male | n.a. | 362,169 (51.5%) | 727,623 (52.1%) | 1,089,792 (51.9%) |
| Background | n.a. | | | |
| Nordic | n.a. | 469,027 (66.6%) | 941,988 (67.5%) | 1411,015 (67.2%) |
| Non-Nordic | n.a. | 234,789 (33.4%) | 454,384 (32.5%) | 689,173 (32.8%) |

* Statistics Norway's country background in three generations: Non-Nordic if either the person in question, or at least one of parents or grandparents were born in a non-Nordic country. M. East: Middle East; N. Africa: North Africa; N. Am: North America.
† Crowded dwelling (household): number of rooms is lower than the number of residents or one resident lives in one room, and the space for each resident is less than 25 $m^2$.
‡ EU60 (EU standard accounting for composition of household members): disposable equivalence income below 60 per cent of the median income.

Supplemental Table S3. Post hoc robustness analyses of the association between SARS-CoV-2 infections or vaccination and risk of type 1 diabetes in Norway.

| Time of SARS-CoV-2 infection | Exposed† | End of follow-up for T1D | Incidence rate T1D per 100,000 person-years | Adjusted Hazard Ratio (95% CI)§ |
|---|---|---|---|---|
| **Pre vs post omicron*** | SARS-CoV-2 **infection** | **As in primary analysis** | | |
| **Age 0-17 y** | | | | |
| 1 Jan 2020 – 30 Nov 2021 | No | 28 Feb 2022 | 42.0 | 1.00 (Reference) |
| | Yes | 28 Feb 2022 | 58.7 | 1.57 (1.00, 2.47) |
| 1 Dec 2021 – 28 Feb 2022 | No | 28 Feb 2022 | 42.0 | 1.00 (Reference) |
| | Yes | 28 Feb 2022 | 71.1 | 1.42 (0.76, 2.67) |
| 1 Jan 2020 – 28 Feb 2022 | No | 28 Feb 2022 | | 1.00 (Reference) |
| | Yes | 28 Feb 2022 | | 1.57 (1.07, 2.29)† |
| | | **Longer follow-up** | | |
| 1 Jan 2020 – 30 Nov 2021 | No | 30 Apr 2024 | 42.9 | 1.00 (Reference) |
| | Yes | 30 Apr 2024 | 49.0 | 1.13 (0.88, 1.45) |
| 1 Dec 2021 – 28 Feb 2022 | No | 30 Apr 2024 | 42.9 | 1.00 (Reference) |
| | Yes | 30 Apr 2024 | 48.9 | 0.95 (0.83, 1.09) |
| 1 Jan 2020 – 28 Feb 2022 | No | 30 Apr 2024 | | 1.00 (Reference) |
| | Yes | 30 Apr 2024 | | 0.99 (0.87, 1.13) |
| | SARS-CoV-2 **vaccination** | | | |
| **Age 12-17 y** | | | | |
| | No | 30 Apr 2024 | 48.9 | 1.00 (Reference) |
| | Yes | 30 Apr 2024 | 47.5 | 1.08 (0.86, 1.37) |
| **Age 18-29** | | | | |
| | No | 30 Apr 2024 | 22.5 | 1.00 (Reference) |
| | Yes | 30 Apr 2024 | 27.8 | 1.06 (0.79, 1.42) |

* Pragmatically dichotomised at 1 December 2021 after which the large majority of infections were omicron variants. † Individuals were counted as exposed from day 30 after positive SARS-CoV-2 test in the specified period or from time of vaccination. ‡ The primary cohort analysis results for Norwegian children aged 0-17 years, shown for comparison. § Adjusted as in the primary results for: age, sex and country background, coeliac disease, family income, crowding, household size, urban/rural residence and geographical region and SARS-CoV-2 vaccination. T1D: Type 1 diabetes. CI: Confidence interval.

Supplemental Table S4. Literature overview: Cohort studies of SARS-CoV-2 infection and risk of type 1 diabetes (age <30 years)

| Study (first author, pub date) | Population | Design | Study period | Age group(s) | Outcome def (diabetes) | Look-back period† | Exposure definition | Tot sample size | n T1D | **Relative Risk*** | Comment |
|---|---|---|---|---|---|---|---|---|---|---|---|
| Kompaniyets 2022 [49] | US Claims database - HealthVerity | Matched cohort (exposed vs non-exp. 1:3) | 1 Mar 2020-31 Jan 2022 | <18 yr | >= entry of T1D code | 1 yr | 60-365 d after PCR or clinical COVID-19 diagnosis | ~3.1 mill | 2872 | 1.23 (1.13-1.33) | T1D incidence very high >120 per 100,000 person-years. Similar to Barrett[60] |
| McKeigue 2022 [46] | All population of Scotland | Registries cohort | Mar, 2020 – 22 Nov 2021 | <35 yr (+subgr <16) | Validated clinical T1D diagnosis in diabetes registry | >=5 years | >=30 d after PCR + vs not | 1,85 mill | 635 | HR 0.86 (0.62-1.21); <16y: 0.79 (0.50, 1.27). | |
| Pietropaolo 2022[51] | Commercial, global claims database - **TriNetX** | Propensity matched within commercial database | 1 Jan 2020-11 Jun 2021 | 0-18 and 19-30 yr | T1D (?), Electronic claims codes | Unclear | >=1 d after PCR+ or COVID diagnosis | ~548 000 in matched analysis (of total ~4 million) | 122 | 0-18 y: RR 0.87 (0.51-1.47); 19-30y: RR 1.87(1.13-3.11) | Details of database and analysis unclear. |
| Chang 2023 [53] | Claims database – **TriNetX** US | Propensity score-matched (1:1). Test-neg. | Jan, 2020 to Dec 2021 | subgroup analysis of 18-40 yr) | T1D ICD-10 diagnoses (plus other) | Not specified | 30-180 days | ~1.8 mill | 4581 | Age 18-40: HR 2.8 (2.5-3.1) | People who received COVID-19 vaccination excluded |
| Kendall 2022 [52] | Commercial, global claims database – **TriNetX** | Propensity score matched (1:1) cohort | March 2020-Dec 2021 | <19 yr | T1D, Electronic claims codes | Unclear | COVID-19 diag. or SARS-CoV-2 PCR+ vs other resp. infection | ~570 000 | 195 | HR (vs resp inf 6 mo after infection): 1.83 (1.36-2.44) | Details of database unclear. Compared to other respiratory infections. |
| Qeadan 2022 [54] | Commercial (USA) claims database **Cerner** | Propensity score matched cohort | Jan 2019 to Jul2021 | All ages (stratified by age groups) | T1D codes (E10.x), but also included O24.x (GDM) | Unclear | >=1 d after exposure | 27.3 million | Age 6-12y: 2354, age 18-35y: >10000 | Age 6-12: OR 2.04 (1.8-2.3); 18-35yr: 0.97 (0.91-1.04) | Details of database unclear |
| Bull-Otterson 2022 [55] | Commercial (USA) claims database **Cerner** | Matched cohort (1:3) | Mar2020-Oct2021 | 18-64 and >=65 yr | Type 1 diabetes disease code | 1 yr | 30-365 d after PCR+ or clin. COVID-19 diagnosis | ~1 mill | 1556 (18-64 yr) + 654 (>=65 yr) | Age 18-64yr: IRR=1.32 (1.19-1.46) | 26 outcomes studied. Incidence rate of T1D said0.02 per 100 |
| Noorzae 2022 [47] | All population of Denmark | Registries cohort + test negative design | Mar2020-Aug2022 | <18 yr | ICD-10 codes in the national patient register | Unclear | >30 d after SARS-CoV-2 test | ~1.1 mill | 613 | HR 0.85 (0.70-1.04) | |
| Zareini 2023 [48] | All Danish residents | Registries matched cohort | To Dec 31, 2021 | <30 yr | ICD-10 codes in national patient-, prescription reg. | Not specified | After SARS-CoV-2 test. from day 31 | ~1.3 mill | 130 | HR 0.69 (0.42-1.13) | Partly overlap w/ Noorzae 2022 |
| Weiss 2023 [56] | Bavarian BASHIP claims | Cohort | 2020 to 2021 | Children, 4-12 yr | ICD-10 codes | Not specified | Not specified | ~1.2 mill | Not specified | HR 1.57 (1.32-1.88) | Not exact dates (quarterly). |

*Measure of association (OR: Odds ratio, HR: Hazard Ratio, RR: Risk ratio, IRR: incidence rate ratio) reporting the ratio of risk (or odds or incidence or hazard) of diabetes among those with versus without exposure to coronavirus or COVID-19. In parenthesis the 95% confidence interval. † Time before incident cases of type 1 diabetes was counted (period for excluding individuals with previous/prevalent diabetes diagnosis). A sufficient period (usually at least one year, often more depending on the health care system) is essential to ensure that the diabetes code/entry/claim used in the analysis represent a truly incident rather than prevalent case of diabetes. For instance, if a patient is in contact with the health care system once every year, then more than one year is necessary to rule out prevalent diabetes. Also, immigration or patients moving between provider/hospital/care provider catchment areas may appear as incident cases unless this is handled in the data base.

Supplemental Table S5. Literature overview: case reports of type 1 diabetes after SARS-CoV-2 vaccination.

| First author, year | Country | Outcome | Vaccine | Dose | Time since vaccination | Age | Sex | Ketoacidosis | GAD-AAb | IA-2-AAb | ICA | Insulin-AAb | ZnT8-AAb | T1D HLA | Preexisting immune conditions/diabetes |
|---|---|---|---|---|---|---|---|---|---|---|---|---|---|---|---|
| Sakurai 2022[61] | Japan | T1D | Cominarty, BNT162b2, Pfizer-BioNTech | 1st dose | 3 days | 36 | Female | Yes | 0 | 0 | n.a | 0 | 0 | Yes | None reported |
| Sasaki 2022[62] | Japan | T1D | Spikevax, mRNA-1273, Moderna | 2nd dose | 4 - 8(?) weeks | 73 | Female | No | 1 | 0 | 0 | 1 | 0 | Yes | Patients HbA1c was >7% 3 months prior to vaccination |
| Sasaki 2022[63] | Japan | Fulminant T1D | Cominarty, BNT162b2, Pfizer-BioNTech | 1st dose | 8 days | 45 | Female | Yes | 0 | 0 | 0 | 0 | 0 | Yes | Stable bronchial asthma |
| Yano 2022[64] | Japan | T1D | Spikevax, mRNA-1273, Moderna | 1st/2nd dose | 40 days since dose 1, 10 days since dose 2 | 51 | Female | Yes | 0 | 0 | n.a | 1 | 0 | Yes | Autoimmunity against thyroid gland at diagnosis |
| Tang 2022[65] | China | Fulminant T1D | CoronaVac, Sinovac | 1st dose | 6 days | 50 | male | Yes | 0 | 0 | n.a | n.a | 0 | Yes | None reported |
| Sato 2022[66] | Japan | ICI–associated fulminant T1D | mRNA vaccine (not specified) | 2nd dose | 14 days | 43 | male | «Ketosis» | 0 | 0 | n.a | n.a | 0 | No | Malignant melanoma, receiving nivolumab treatment |
| Bleve 2022[67] | Italy | T1D | Vaxzevria, ChAdOx1-S, Oxford-AstraZeneca | 1st dose | 7 days | 57 | Female | yes(?) | 1 | 1 | n.a | n.a | n.a | n.a | Anti-Tg2 antibodies positive at diagnosis |
| | | | Cominarty, BNT162b2, Pfizer-BioNTech | 1st dose | 24 days | 61 | Female | n.a (0?) | 1 | n.a | n.a | n.a | n.a | n.a | Acquired hypothyroidism |
| Patricio 2021[68] | Italy | T1D and Graves disease | Cominarty, BNT162b2, Pfizer-BioNTech | 2nd dose | 4 weeks | 52 | Male | n.a (0?) | 1 | n.a | n.a | n.a | n.a | n.a | Vitiligo and T2D |

T1D – Type 1 Diabetes; ICI – Immune Checkpoint Inhibitor; HLA – Human Leukocyte Antigen (Yes: patient had a susceptibility variant)

### Supplemental Figures

#### Supplemental Figure S1. SARS-CoV-2 infections and vaccinations over time by age in Norway and Sweden

**Norway** **Sweden**

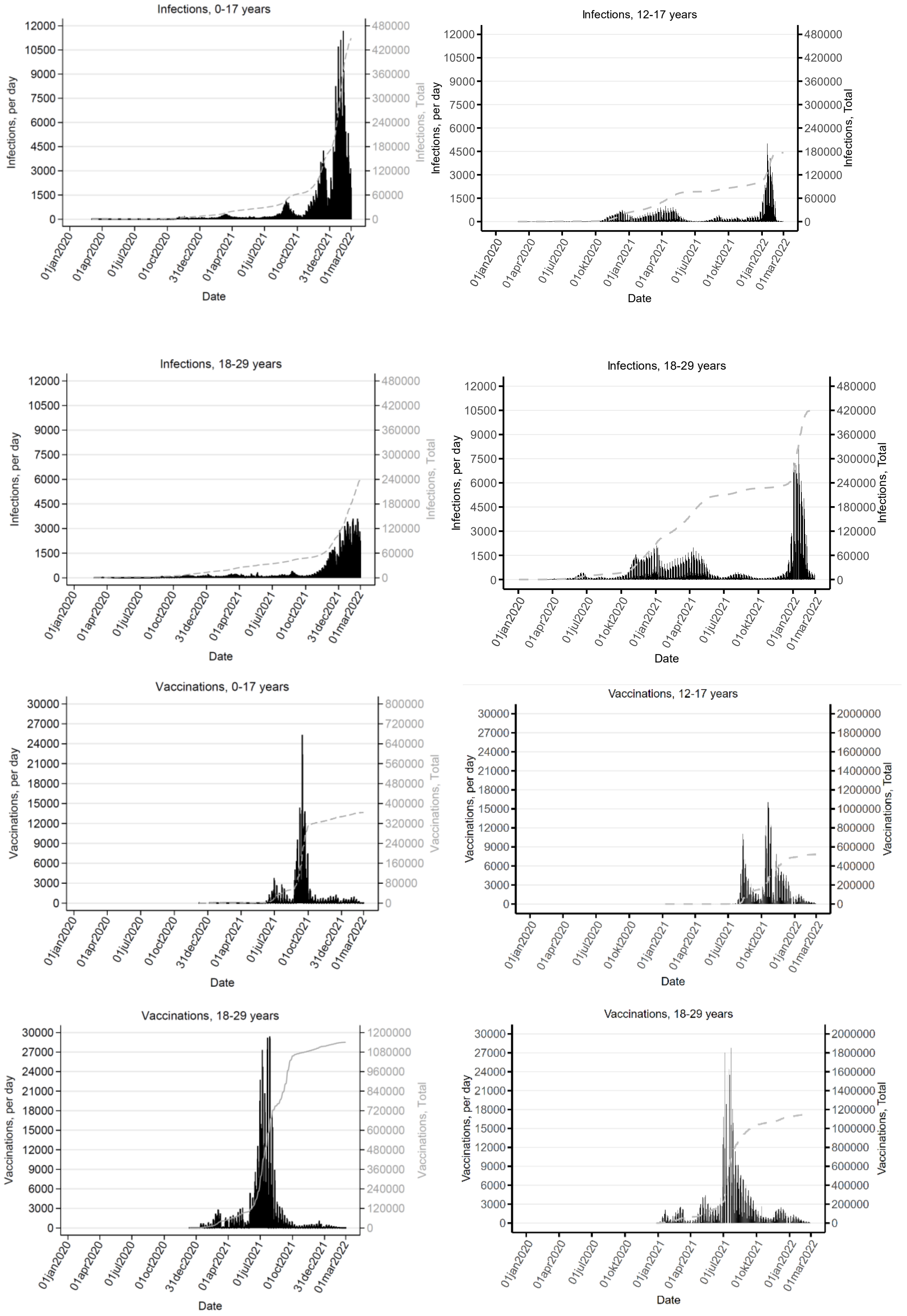

Supplemental Figure S2. Distribution of follow-up time from SARS-CoV-2 infection or vaccination to newly diagnosed type 1 diabetes in Norway. a: Age 0-17 years (infection in upper panel and vaccination in lower panel). b: Age 18-29 years (infection in upper panel and vaccination in lower panel). In the association analyses shown in Main Figure 2, only time from day 31 after infection or vaccination was counted as exposed time.

a

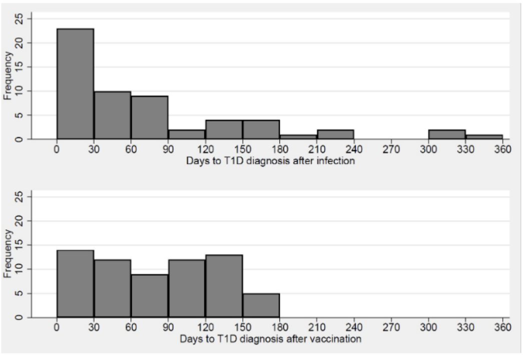

b

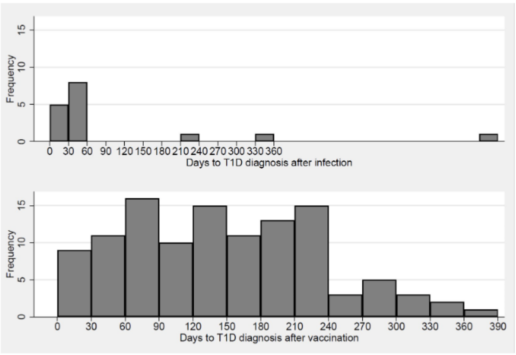

Supplemental Figure S3. Distribution of times from SARS-CoV-2 infection or vaccination to newly diagnosed type 1 diabetes in Sweden.

a: days from SARS-CoV-2 infection to type 1 diabetes. b: Days from SARS-CoV-2 vaccine to diagnosis of type 1 diabetes. The first 180 days of follow-up shown here for the age-group 12-29 years. In the association analyses shown in Main Figure 2, only time from day 31 after infection or vaccination was counted as exposed time.

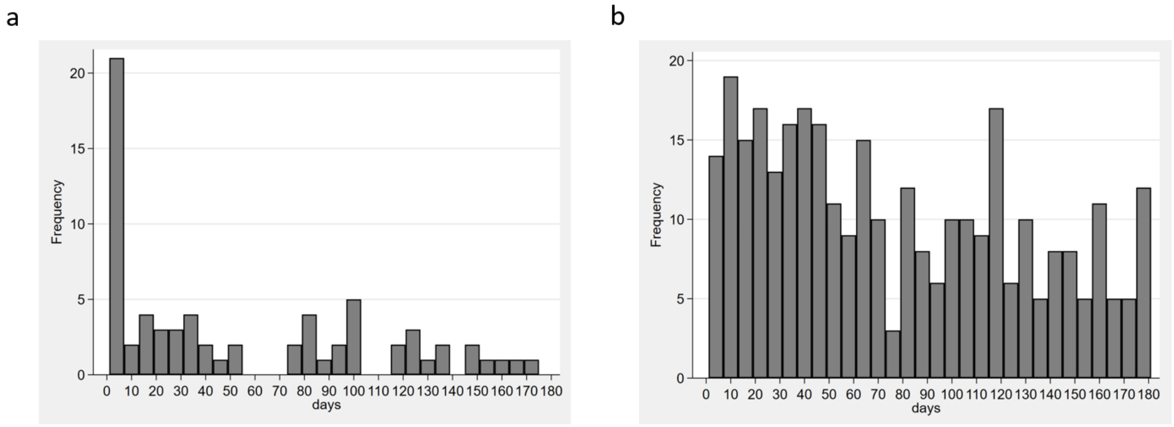

Supplemental Figure S4. Additional results for association between SARS-CoV-2 infections and risk of type 1 diabetes in Norway: age-group 0-17 years and results from test-negative design analysis.

These are results to supplement Main Figure 2a. Cohort analysis results for age 12-17 and 18-29 years are the same as shown in Main Figure 2a. Test neg.: Test-negative design analysis where test positive are compared to test negatives and those never tested for SARS-CoV-2 infection are excluded from the analysis.

| Country | Age (years) | # T1D in exp./unexp. | IR/100,000 PYR in exp./unexp. | Analysis | Adjusted Rate Ratio (95% CI) |
|---|---|---|---|---|---|
| Norway | 0-17 | 35/278 | 59.0/40.4 | Test neg. | 1.57 (1.07, 2.29) |
| Norway | 0-17 | 35/916 | 59.2/41.9 | Cohort | 1.49 (1.03, 2.14) |
| Norway | 12-17 | 19/156 | 57.2/42.8 | Test neg. | 1.58 (0.93, 2.70) |
| Norway | 12-17 | 18/331 | 63.1/43.0 | Cohort | 1.80 (1.07, 3.02) |
| Norway | 18-29 | 11/168 | 22.3/24.3 | Test neg. | 0.80 (0.43, 1.47) |
| Norway | 18-29 | 11/345 | 22.3/21.1 | Cohort | 0.83 (0.45, 1.52) |

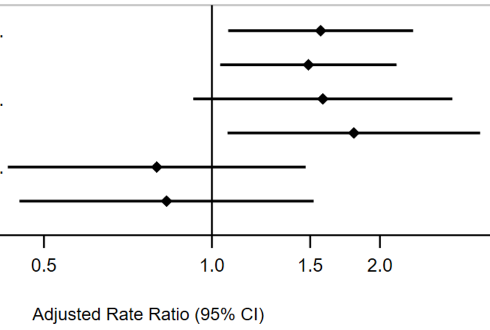

Supplemental Figure S5. Adjusted and unadjusted rate ratios for associations of SARS-CoV-2 infections and vaccinations with risk of type 1 diabetes. a: infections. b: vaccinations. Horizontal bars are 95% confidence intervals (CI). Adjusted data are the same as shown in Main figure 2. Analyses were adjusted for age, sex and Nordic or non-Nordic country background. Additional adjustments for coeliac disease, family income, crowding, household size, urban/rural residence and geographical regions were done for Norwegian analyses (see Supplemental statistical analysis section above for additional details).

a

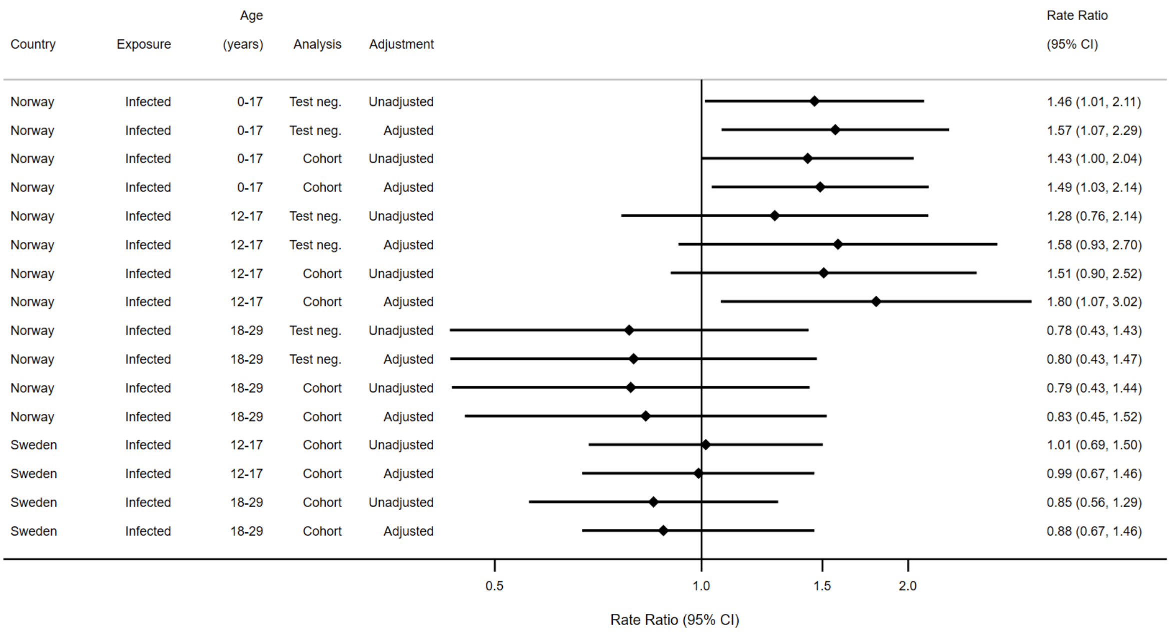

b

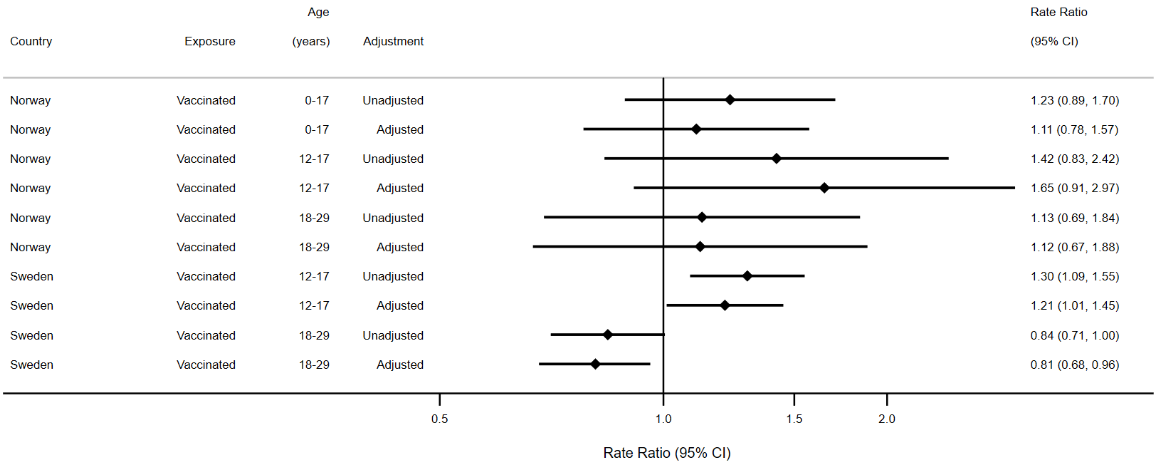

Supplemental Figure S6. Post hoc association analysis of SARS-CoV-2 vaccination dose 1 and 2.

Adjusted rate ratios for type 1 diabetes after one dose (a) or two doses (b) vs no dose of SARS-CoV-2 mRNA vaccine in age-groups 12-17 years and 18-29 years in Norway and Sweden. Follow-up time counted only from the 31st day after the first dose (a) or second (or subsequent) dose (b). Vaccine doses modelled as time-varying exposure. Adjustment for covariates as in Main Figure 2.

a

| Country | Age (years) | # T1D in exp./unexp. | IR/100,000 PYR in exp./unexp. | Exposure | Adjusted Rate Ratio (95% CI) |
|---|---|---|---|---|---|
| Norway | | | | | |
| Norway | 12-17 | 40/291 | 50.7/42.9 | Vaccinated, 1st dose vs none | 1.69 (0.92, 3.09) |
| Norway | 18-29 | 16/244 | 28.8/19.6 | Vaccinated, 1st dose vs none | 1.30 (0.67, 2.53) |
| Subgroup, DL ($I^2$ = 0.0%, p = 0.569) | | | | | 1.50 (0.96, 2.35) |
| Sweden | | | | | |
| Sweden | 12-17 | 15/330 | 37.6/38.6 | Vaccinated, 1st dose vs none | 0.94 (0.55, 1.60) |
| Sweden | 18-29 | 26/278 | 32.6/27.5 | Vaccinated, 1st dose vs none | 1.16 (0.78, 1.73) |
| Subgroup, DL ($I^2$ = 0.0%, p = 0.536) | | | | | 1.08 (0.78, 1.48) |
| Heterogeneity between groups: p = 0.237 | | | | | |
| Overall, DL ($I^2$ = 0.0%, p = 0.551) | | | | | 1.20 (0.93, 1.56) |

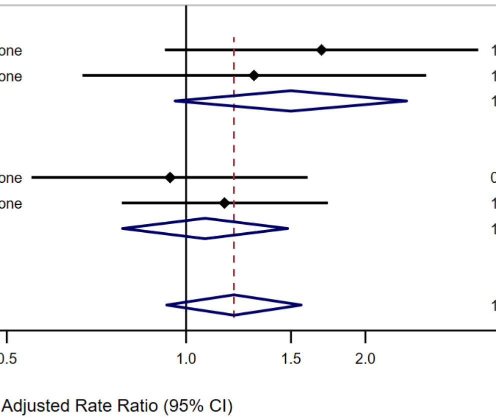

b

| Country | Age (years) | # T1D in exp./unexp. | IR/100,000 PYR in exp./unexp. | Exposure | Adjusted Rate Ratio (95% CI) |
|---|---|---|---|---|---|
| Norway | | | | | |
| Norway | 12-17 | 7/291 | 38.0/42.9 | Vaccinated, 2nd dose vs none | 1.56 (0.60, 4.07) |
| Norway | 18-29 | 65/244 | 26.7/19.6 | Vaccinated, 2nd dose vs none | 1.20 (0.67, 2.15) |
| Subgroup, DL ($I^2$ = 0.0%, p = 0.648) | | | | | 1.29 (0.78, 2.12) |
| Sweden | | | | | |
| Sweden | 12-17 | 176/330 | 51.3/38.6 | Vaccinated, 2nd dose vs none | 1.24 (1.02, 1.49) |
| Sweden | 18-29 | 142/278 | 23.8/27.5 | Vaccinated, 2nd dose vs none | 0.83 (0.68, 1.01) |
| Subgroup, DL ($I^2$ = 87.9%, p = 0.004) | | | | | 1.02 (0.69, 1.51) |
| Heterogeneity between groups: p = 0.464 | | | | | |
| Overall, DL ($I^2$ = 67.5%, p = 0.027) | | | | | 1.08 (0.80, 1.45) |

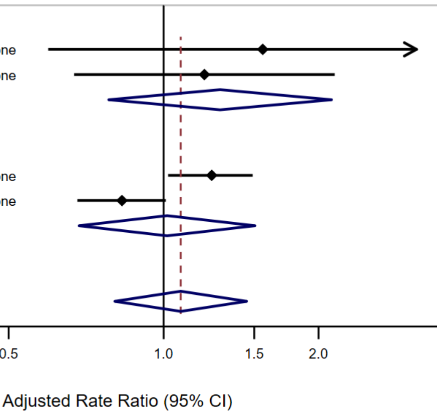

Supplemental Figure S7. Supplemental and analyses of severity at onset of type 1 diabetes.

Trends in mean blood pH, serum bicarbonate at diagnosis of type 1 diabetes during 2016-2023 in Norway. a and b are scatterplots with predictions from linear regression with splines shown as dashed dark blue line. c: Trends in moderate and d: severe DKA with shaded 95% CI and predictions from linear interrupted time series logistic regression models (dashed purple line). There were no significant time trends or break in trend for moderate or severe DKA according to the interrupted time series analysis. Moderate DKA was defined as pH <7.2 or serum bicarbonate <10 mmol/L, and severe DKA was defined as pH <7.1 or serum bicarbonate <5 mmol/L. T1D: Type 1 diabetes; DKA: Diabetic ketoacidosis; CI: confidence interval.

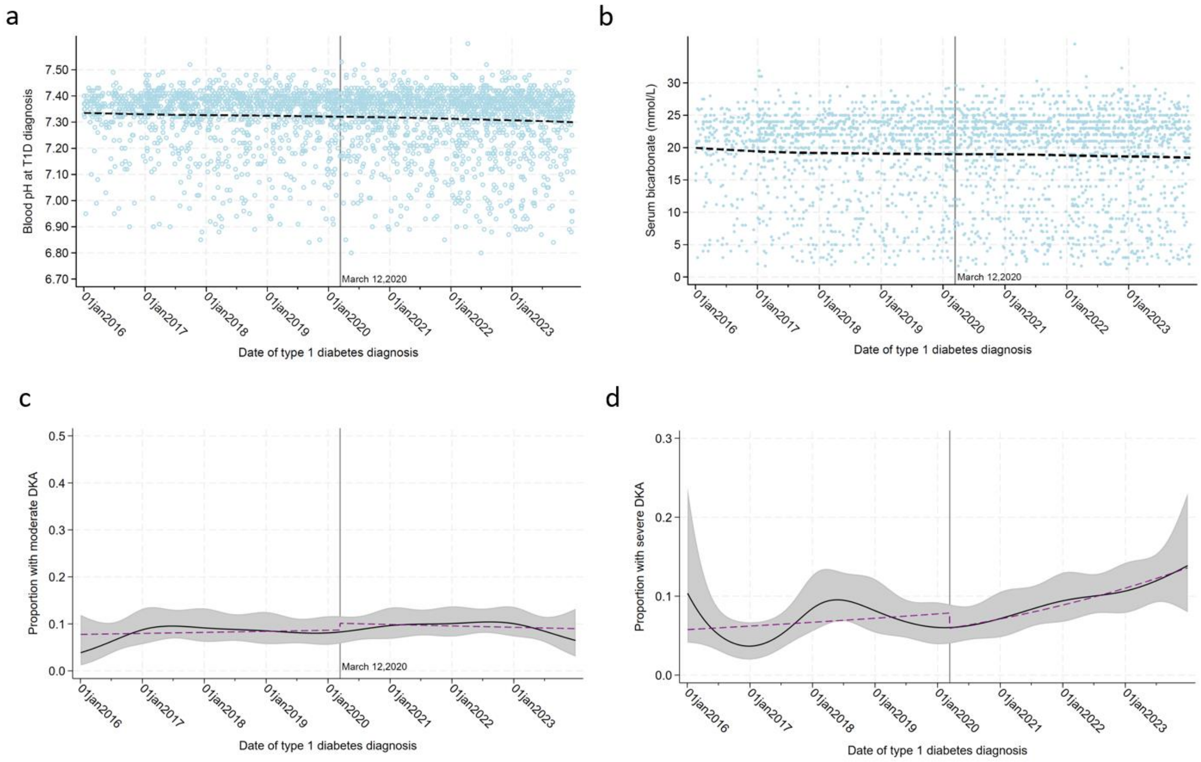

Supplemental Figure S8. Forest plot of infection estimates from previous studies and the current study. Estimates are relative risks or rates (RR) for the association between SARS-CoV-2 infections and risk of type 1 diabetes, subdivided by age-group. Details of previous cohort studies shown in Supplemental Table S3, and search and selection strategy described in literature review section above. References listed in Supplemental references: Kompaniyets 2022 [49], McKeigue 2022 [46], Pietropaolo 2022[51], Chang 2023 [53], Kendall 2022 [52], Qeadan 2022 [54], Bull-Otterson 2022 [55], Noorzae 2022 [47], Zareini 2023 [48], Weiss 2023 [56]. Where database is not mention, the data came from individually linked nation-wide population based studies similar to the current study. Results shown are from age-groups most similar to those in the current study. Pooled estimates were not produced because of heterogeneity in study design and potential overlap in data form the same sources.

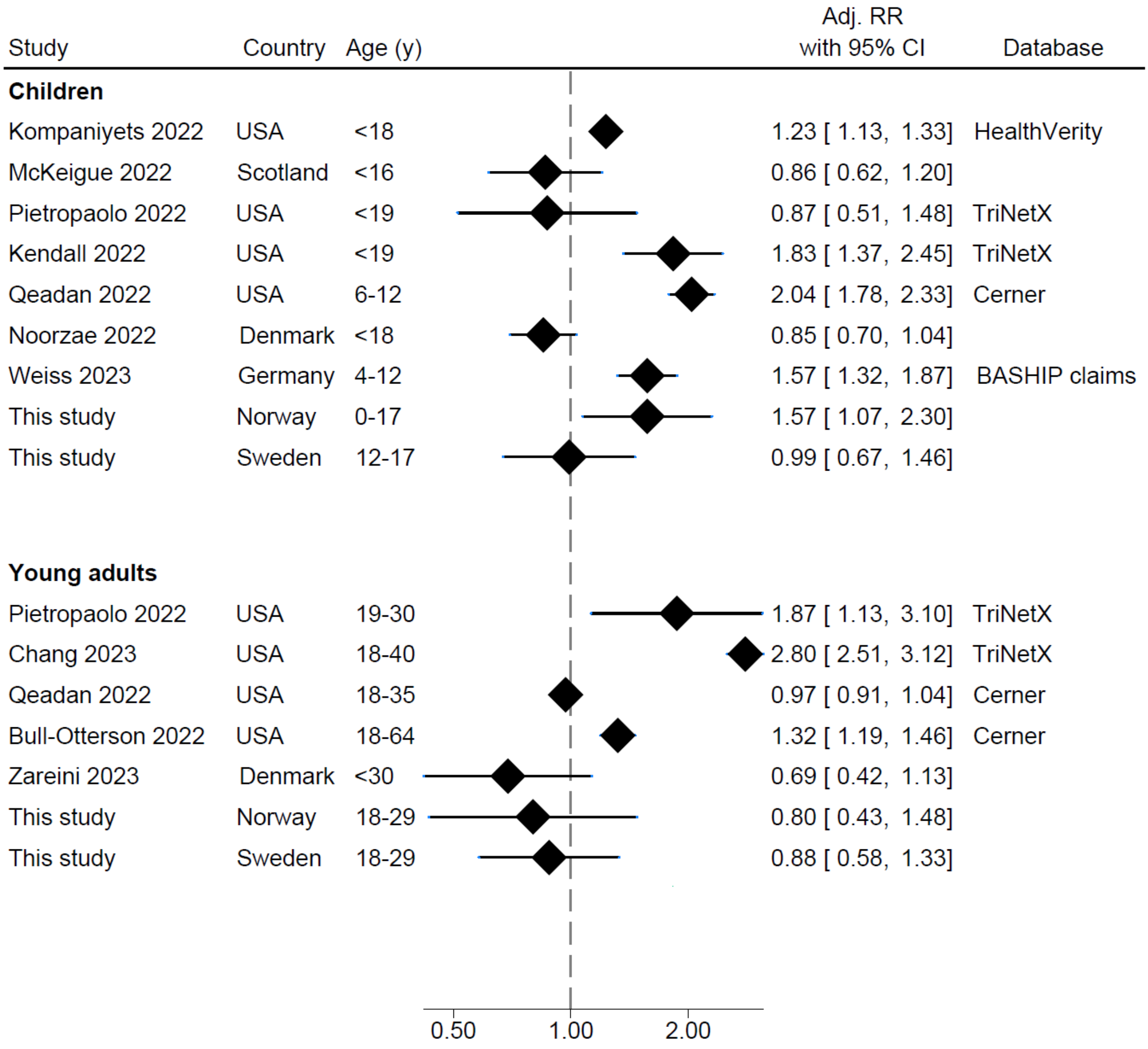
